# Lockdown benefit varies among countries and sub-national units: a reanalysis of the data by Bendavid et al. (2021)

**DOI:** 10.1101/2021.02.17.21251898

**Authors:** Lukas Boesch

## Abstract

Are the lockdown measures limiting the propagation of COVID-19? Recent analyses on the effectiveness of non-pharmaceutical interventions in reducing COVID-19 growth rates delivered conflicting conclusions. While Haug et al. (2020) did find strong empirical support for reductions in COVID-19 growth rates, Bendavid et al. (2021) did not. Here, I present the results of a reanalysis of the data by Bendavid et al. (2021). Instead of relying on pairwise comparisons between 10 countries with fixed-effects regression models to isolate the effect of lockdown measures, I modelled the development of the pandemic with and without lockdown measures for the entire period and all countries included in the data with one mixed-effects regression model. My results reconciled the conflicting conclusions of Haug et al. (2020) and Bendavid et al. (2021): while mandatory business closure orders did not affect COVID-19 growth rates, a general decrease in COVID-19 growth rates was attributable to the implementation of mandatory stay-at-home orders. However, the effect of mandatory stay-at-home orders varied, being weaker, even zero, in some countries and sub-national units and stronger in others, where COVID-19 growth rates only decreased due to the implementation of mandatory stay-at-home orders. The heterogeneity in the effect of mandatory stay-at-home orders on the spread of COVID-19 is challenging from a scientific and political point of view.

## Introduction

Are non-pharmaceutical interventions (NPIs) helping to limit the spread of COVID-19 infections? Most importantly, are lockdown measures, the more restrictive NPIs (mrNPIs) based on mandatory stay-at-home and business closure orders, efficient in reducing COVID-19 growth rates? Two recently published analysis resulted in contradicting evidence. On the one hand, Haug et al. (2020) concluded that mrNPIs were the most effective NPIs but showed a considerable variation in effectiveness across countries. On the other hand, Bendavid et al. (2021) denied the possibility of large declines in COVID-19 growth rates due to mrNPIs: “While modest decrease in daily growth (under 30%) cannot be excluded, the possibility of large decreases in daily growth due to mrNPIs is incompatible with the accumulated data.” (2021, p. 8).

In this paper, I present a reanalysis of the data used by Bendavid et al. (2021). I run one mixed-effects regression that compared the daily changes in COVID-19 growth rates across all countries and sub-national units included in the data to model the development of the pandemic with and without the implementation of mrNPIs. I believe that this approach provides a more accurate assessment of the effectiveness of mrNPIs in decreasing COVID-19 growth rates than the one taken in Bendavid et al. (2021), which relied on the unrealistic assumption that Sweden and South Korea are counterfactuals of England, France, Germany, Iran, Italy, the Netherlands, Spain, and the United States.

## Methods

Bendavid et al. (2021) used an analytical framework that was based on the assumption that countries that did not implement mrNPIs were counterfactuals to countries that did implement mrNPIs: „Here, we use Sweden and South Korea as the counterfactuals to isolate the effects of mrNPIs in countries that implemented mrNPIs” (Bendavid et al. 2021, p. 5). Based on this assumption, the authors run pairwise fixed-effects regression models to compare the combined effect size of all NPIs on the daily COVID-19 growth rates of countries that did implement mrNPIs (the treatment countries England, France, Germany, Iran, Italy, the Netherlands, Spain, and the United States) with the combined effect size on the daily COVID-19 growth rate of all NPIs of countries that did not implement mrNPIs (the control countries South Korea and Sweden). The analysis was based on a time-series of COVID-19 case counts at the sub-national unit level, matched with data on the implementation of a variety of NPIs during winter/spring 2020. The dependent variable was the daily difference in the natural log of the number of cumulated COVID-19 cases. Only cumulated daily differences equal or larger ten were considered.

The authors claimed that their approach allowed to isolate the effect of mrNPIs. A closer look, however, revealed that this was a misleading view. First, the classification of countries and their sub-national units either as control or treatment ignored the huge differences in the implementation of the two mrNPIs. For example, only three treatment countries implemented mandatory business closure, and this with varying intensity. Furthermore, while seven sub-national units in the USA did not implement mandatory home isolation at all, only the minority of French and Italian sub-national units did so with maximal intensity. On the other hand, all German and Dutch regions implemented mandatory home isolation with full intensity (appendices 1 and 2). Second, this approach did not tease apart the effects of business closure and home isolation, with the risk of drawing the wrong conclusion: for example, if one is positive and the other negative, the combined effect is zero. Finally, the treatment and control countries are not counterfactuals. Even if the additional benefit of implementing mrNPIs was neglectable in a direct comparison of countries, we still not know how daily COVID-19 growth rates would have evolved if the countries had/had not implemented mrNPIs.

I solved these issues by modelling the counterfactual outcomes. Based on the data provided by Bendavid et al. 2021, I built one dataset that included all observations of all sub-national units of all ten countries of interest for the entire period available. In order to model a dynamic effect of the mrNPIs on the daily growth rate in COVID-19 cases, I generated the variable home, which was the cumulated sum of days after a sub-national unit implemented home isolation, weighted with the intensity of implementation, and the variable busi, which was the cumulated sum of days after a sub-national unit implemented business closure, weighted with the intensity of implementation. To control for the unobserved heterogeneity and model the time trend, I computed the variable days, which simply was a count of the epidemic age, in days, after a sub-national unit had reached cumulative confirmed cases of 10. Then, I narrowed down the extent of the time series such as to include the same cases as in Bendavid et al. (2021). Finally, I log transformed all three variables (home and busi after the addition of one) and generated the cluster means for epidemic age (mdaysc and mdayss), for weighted days with home isolation (mhomec and mhomes) and for weighted days with business closures (mbusic and mbusis). In total, the data set used for the reanalysis included 5324 observations, clustered within 10 countries and 209 sub-national units. The dates of the time series spanned from 18.02.2020 to 06.04.2020. The number of observations at the sub-national units level ranged from three to 49 (mean=25.47; sd=8.33). Epidemic age ranged from one to 67 days (mean=15.71; sd=10.38) Weighted home isolation days from zero to 29 (mean=4.84; sd=6.83) and weighted business closure days from zero to 32.75 (mean=2.87; sd=5.56) (see appendix 3 for detailed summary statistics).

I modelled the daily growth rate of COVID-19 cases, g_cs_, as a function of a deviation from an overall mean B_0_. The overall mean varied across countries (country random intercept RI_0c_) and sub-national units (units random intercept RI_0s_). Furthermore, g_cs_ was also determined by an overall time trend, B_0days_, which also varied across countries (country random slope RS_cdays_) and sub-national units (units random slope RS_sdays_). These 6 terms captured the entire variation in g_cs_ due to the countries, the sub-national units and their specific time trends that also included the combined effects of all implemented NPIs (excluding home isolation and business closure). This was the baseline model. By adding a fixed effect for business closure (B_0busi_) and a fixed effect as well as random slopes for home isolation (B_0home_; RS_chome_; RS_shome_), the effect of home isolation and business closure on the change in daily COVID-19 growth rates was estimated, while controlling the country and sub-national unit specific time trend. I did not include random slopes for B_0busi_ as only the minority of countries and sub-national units implemented business closure. Finally, I also included the cluster specific means of the predictor variables into the model to account for the correlation between the random intercepts and the regressors and avoid violating the exogeneity assumption, as proposed in Antonakis et al. (2019). This was the full model (equation 1). I used the estimates of the full model to predict the evolution of daily COVID-19 growth rates for the combined sample data, for each country and for a set of sub-national units seperately in both counterfactual conditions (no implementation of mrNPIs vs. maximal implementation of mrNPIs).

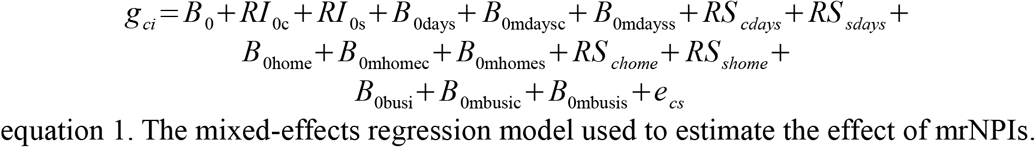

The model in equation 1 was estimated with the glmer function of the lme4 package (Bates et al. 2015) in R (R Core Team 2019). I used a gamma error distribution with a log link to keep estimated values in the range of defined values (exclusion of negative values) and to avoid violating distributional assumptions made for a model with Gaussian error distribution. Prior to estimation, growth rates equal to zero were replaced with the smallest growth rate observed, divided by ten (6.78196e-05). All predictor variables were standardized to a a standard deviation of one and a mean of zero prior to estimation to achieve easier interpretable estimates (Schielzeth 2010). P-values for the fixed effects were obtained by comparing the fit of the full model with the fit of reduced models excluding single fixed effects. P-values for the random slopes were computed by comparing the fit of the full model with the fit of models excluding single random slopes and dividing the resulting p-values by two, as proposed in Bolker et al. (2009). The multicollinearity among regressors was assessed by applying the vif function of the car package (Fox & Weisberg 2019) to a multiple linear regression model that only included the fixed effects from the full model (appendices 4 and 5). I ran a model sensitivity analysis, excluding single countries, to check for robustness of estimates (appendix 6). I checked for violations of distributional assumptions of the random terms with histograms (appendix 7) and checked for overdispersion with an overdispersion test (appendix 8). Inferences on the difference in trajectories of the counterfactual outcomes were based on bootstrapped model estimates.

## Results

While both estimates for home isolation and business closure were negative, only the estimate for home isolation was significant as well as robust to the exclusion of countries (appendix 6 and appendix 9). All random slopes, as well as the fixed effect for epidemic age, were significant (appendix 9). Overall, COVID-19 growth rates decreased with increasing epidemic age. However, the model estimates also showed that the decrease in COVID-19 growth rates was stronger when mandatory home isolation was implemented. The implementation of mandatory business closure, on the other hand, was not associated with an additional decrease in COVID-19 growth rates (figure 1). Interestingly, the effect of home isolation was not the same across all countries. While the model predictions for the counterfactual control outcome (wihout home isolation) did not differ substantially from the model predictions for the counterfactual treatment outcome (with home isolation) in four countries (above South Korea and Sweden this was also the case for Iran and Italy), the model predictions suggested that home isolation had a clear negative effect on COVID-19 growth rates in five countries. For England, the effect of home isolation was uncertain (figure 2). Finally, the effect of home isolation did also vary among sub-national units. In Spain, for example, the negative effect of home isolation on the daily COVID-19 growth rates was not generalizable to all regions. In Navarra, Extremadura and Murcia, the predicted effect of home-isolation was null. In Catalonia and four other regions, on the other hand, model predictions suggested that COVID-19 growth rates would have increased, and not decreased, without the implementation of home isolation (figure 3).

**Figure 1.**
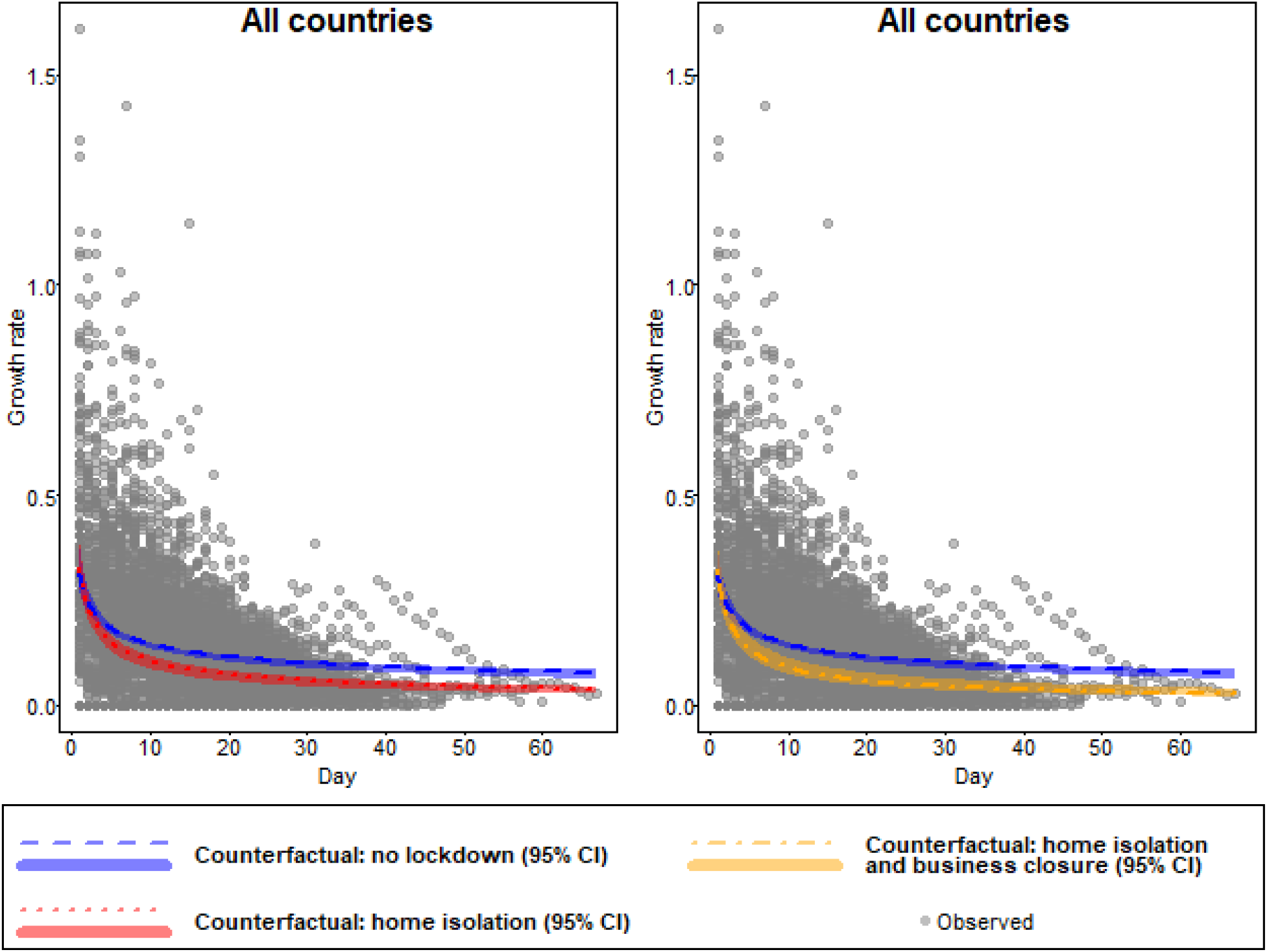
Modelled counterfactual daily COVID-19 growth rates for control (no implementation of home isolation) and treatment (implementation of home-isolation and business closure from the beginning and with full intensity) for all countries and for the entire period of the time series. The prediction for the counterfactual control outcome was calculated by building a sequence of days covering the entire period of the time-series, multiplying each value of this sequence with the fixed effect of epidemic age and adding the population intercept. The prediction for the counterfactual treatment outcome with home isolation was calculated by adding the product between the sequence of days and the fixed effect of home isolation to the values of the counterfactual control outcome. The prediction for the counterfactual treatment outcome with home isolation and business closure was calculated by adding the product between the sequence of days and the fixed effect of business closure to the values of the counterfactual treatment outcome with home isolation. 95% confidence intervals were computed based on the predictions of 1000 bootstrapped models (the R-function was provided by Roger Mundry).

**Figure 2.**
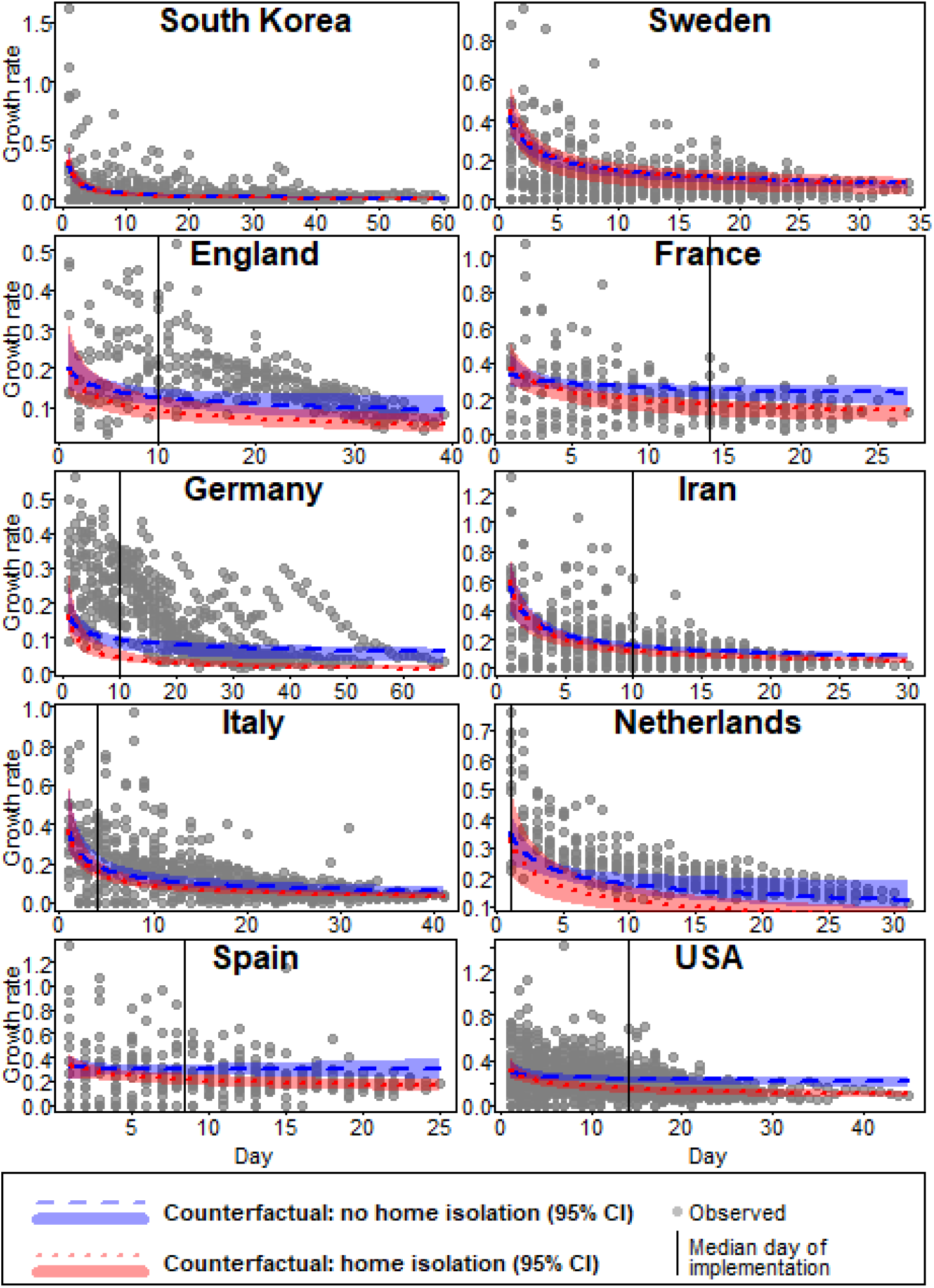
Modelled counterfactual daily COVID-19 growth rates for control (no implementation of home isolation) and treatment (implementation of home-isolation from the beginning and with full intensity), by country and for the entire period of the time series. All model predictions are for average days with business closure. The predictions for the counterfactual control outcome were calculated by building a sequence of days covering the entire period of the time-series of the country and multiplying each value of this sequence with the fixed effect of epidemic age and the country-specific effect of epidemic age. The population intercept, the country-specific intercept as well as the effects of the different population cluster means were added to the resulting values. The predictions for the counterfactual treatment outcome were calculated by adding the product between the sequence of days and the fixed effect of home isolation and the country-specific effect of home isolation to the control outcome. 95% confidence intervals were computed based on the predictions of 1000 bootstrapped models (the R-function was provided by Roger Mundry). For South Korea and Sweden, the model predictions for the counterfactual treatment outcome entirely lacked data support. For all other countries, the model predictions for the counterfactual treatment outcome prior to the implementation of the measure (left of the vertical line) were an extrapolation of the predicted trend after implementation.

**Figure 3.**
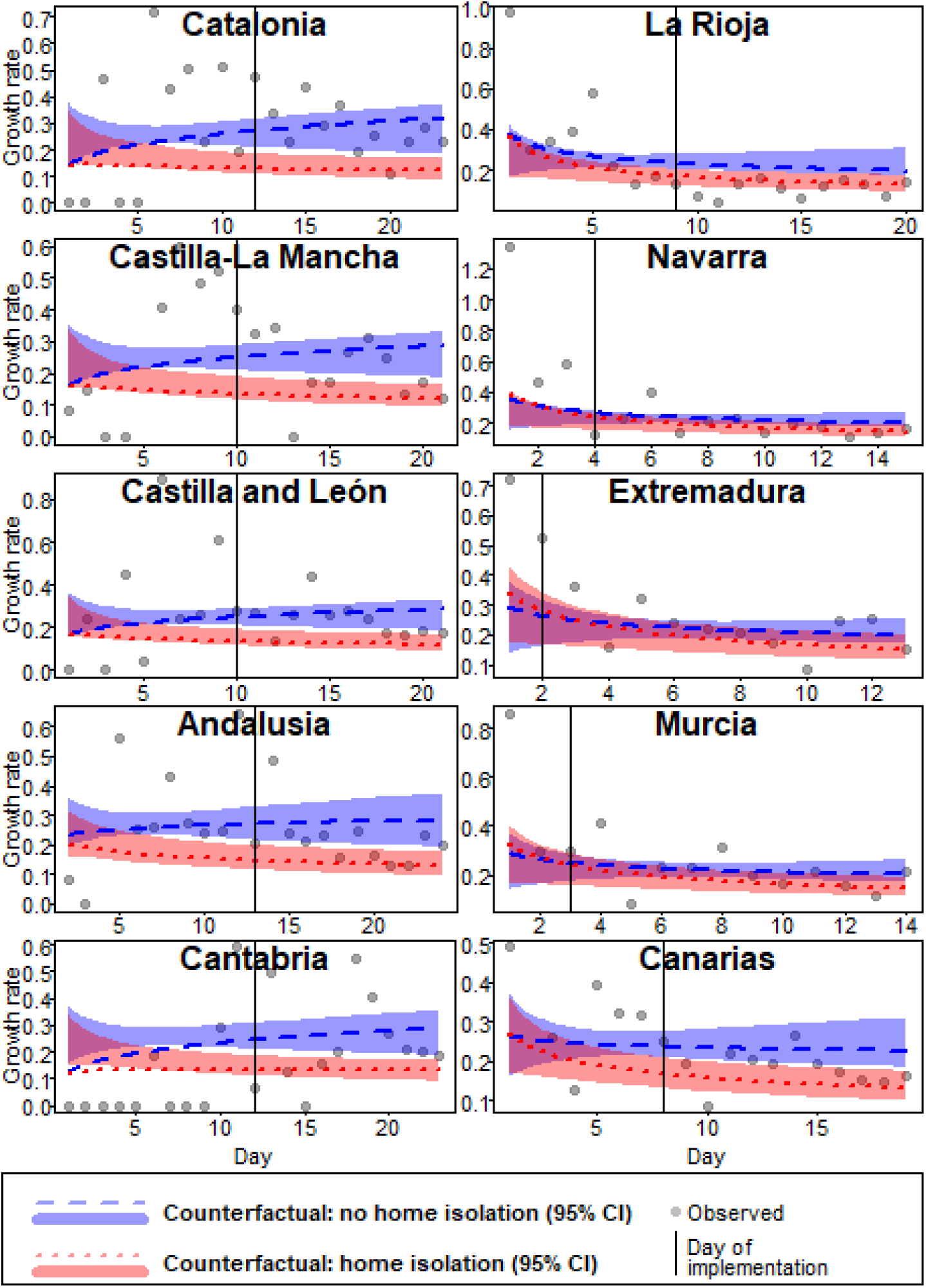
Modelled counterfactual daily COVID-19 growth rates for control (no implementation of home isolation) and treatment (implementation of home-isolation from the beginning and with full intensity), for the entire time series of the ten Spanish regions with the strongest deviations from the country average. All model predictions are for average days with business closure. The predictions for the counterfactual control outcome were calculated by building a sequence of days covering the entire time-series of the region and multiplying each value of this sequence with the fixed effect of epidemic age, the country-specific effect of epidemic age and the region-specific effect of epidemic age. The overall intercept, the country-specific intercept, the region-specific intercept and the effects of the cluster means were added to the resulting values. The predictions for the counterfactual treatment outcome were calculated by adding the product between the sequence of days and the fixed effect of home isolation, the country-specific effect of home isolation and the region-specific effect of home isolation to the values of the control outcome. 95% confidence intervals were computed based on the predictions of 1000 bootstrapped models (the R-function was provided by Roger Mundry). For all regions, the model predictions for the counterfactual treatment outcome prior to the implementation of the measure (left of the vertical line) were an extrapolation of the predicted trend after implementation.

## Conclusion

In this paper, I presented a reanalysis of the data used by Bendavid et al. (2021). On the one hand, I replicated the main finding of the authors: the data do not support the hypothesis that the implementation of mrNPIs leads to a general strong decline in daily COVID-19 growth rates. Although home isolation did lead to a general decline in COVID-19 growth rates, business closure did not affect COVID-19 growth rates. Furthermore, the average decline in COVID-19 growth rate was strong even without the implementaion of home isolation.

On the other hand, the average effect of home isolation showed a statistically significant variation across levels of countries and sub-national units. The inclusion of home isolation random slopes at both levels substantially increased the predictive power of the model. While for some countries and sub-national units, home isolation was necessary to decrease COVID-19 growth rates, in most countries and sub-national units the additional effect of the implementation of home isolation was moderate or zero. The heterogeneity of the effectiveness of NPIs, their interaction with the countries of implementation, was a central finding of the study by Haug et al. (2020). Here, I showed that this heterogeneity was also present in the data by Bendavid et al. (2021) and also applies to the sub-national unit levels.

To some extent, my findings are recomforting, as they allow to reconcile the opposing conclusions by Haug et al. (2020) and Bendavid et al. (2021). However, this reconciliation results from an artificial methodological separation between the average treatment effect, on the one side, and the heterogeneity of the treatment effect, on the other side. From a theoretical point of view, as long as there is a systematic variation of the average treatment effect, which is based on some population clusters, the treatment does have an effect on the outcome. Furthermore, as long as the mechanisms driving the interaction between population clusters and the treatment effect have not been uncovered, it remains unknown how the treatment truly works.

This is the position we are in now. We know that there is no strong average effect of home isolation on the COVID-19 growth rates. However, we also know that home isolation can be highly effective in decreasing daily COVID-19 growth rates in some settings. As long as the mechanisms responsible for the interaction between home isolation and the social context have not been uncovered, it will not be possible to predict where and when home isolation will lead to decreased COVID-19 growth rates. Eventually, this understanding will result from the incorporation of factors not usually considerered in epistemological models, such as complex social networks (Manzo 2020). Moreover, our knowledge is based on a deprecated database. Most countries in the world experience the 2nd or even 3rd wave of the COVID-19 pandemic, with records of implemented NPIs that span periods of up to one year. The first wave is a selective and unrepresentative sample of the epidemic. The assessment of the effectiveness of NPIs must be conducted as soon as possible with an updated database that includes all phases of the pandemic and not only the beginning.

## Supporting information

Functions and script for the analysis

## Data Availability

Data is already available in Bendavid et al. (2021)

https://onlinelibrary.wiley.com/doi/10.1111/eci.13484

## Acknowledgement

I thank Roger Mundry for his valuable advices. Most importantly, he provided the functions for bootstrapping and the overdispersion test. I am very grateful for his support. I also thank Christophe Boesch for reviewing my manuscript, Ivar Krumpal and Valentina Viego for their constructive feedback and Roger Berger for his general support.

## Appendix

1) The implementation and intensity of home isolation, by country.The total column shows the proportion of regions that issue mandatory stay-at-home orders

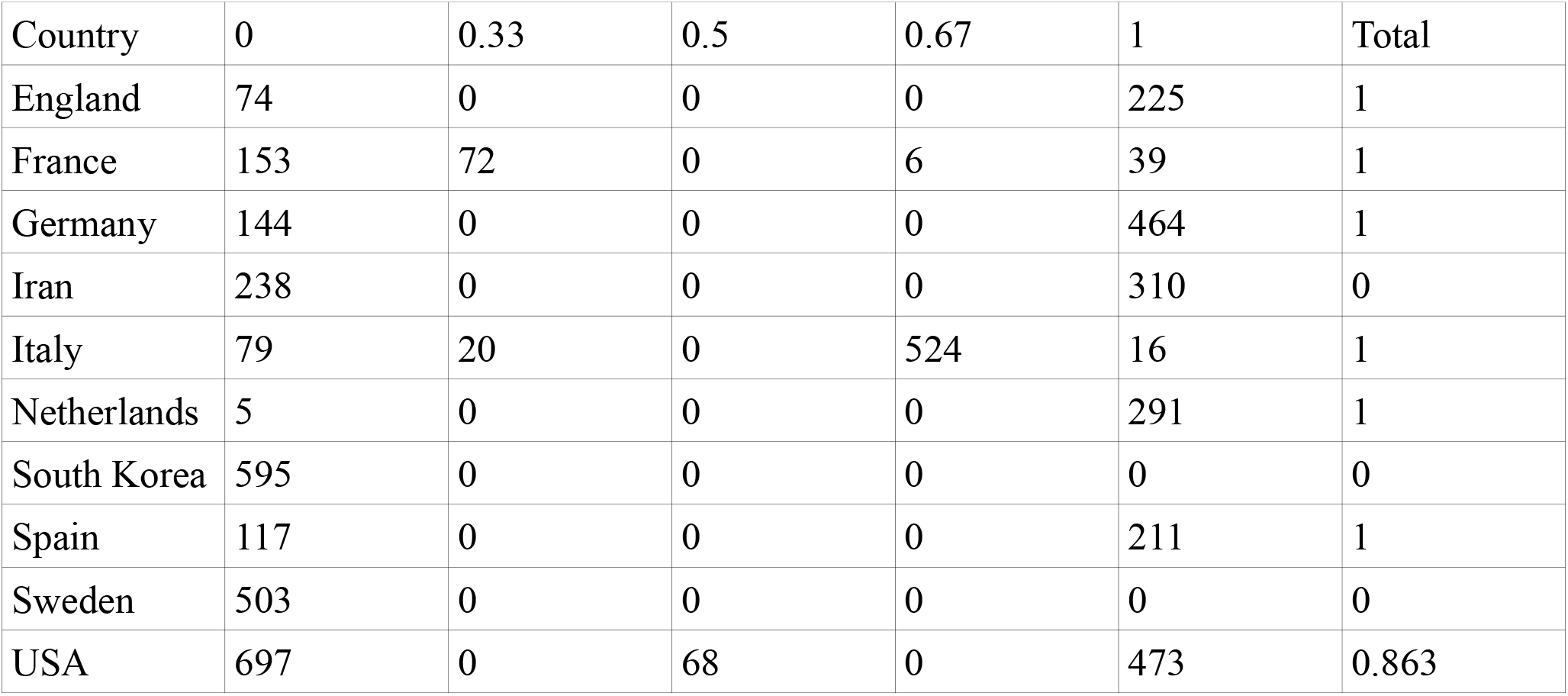

| Country | 0 | 0.33 | 0.5 | 0.67 | 1 | Total |
| --- | --- | --- | --- | --- | --- | --- |
| England | 74 | 0 | 0 | 0 | 225 | 1 |
| France | 153 | 72 | 0 | 6 | 39 | 1 |
| Germany | 144 | 0 | 0 | 0 | 464 | 1 |
| Iran | 238 | 0 | 0 | 0 | 310 | 0 |
| Italy | 79 | 20 | 0 | 524 | 16 | 1 |
| Netherlands | 5 | 0 | 0 | 0 | 291 | 1 |
| South Korea | 595 | 0 | 0 | 0 | 0 | 0 |
| Spain | 117 | 0 | 0 | 0 | 211 | 1 |
| Sweden | 503 | 0 | 0 | 0 | 0 | 0 |
| USA | 697 | 0 | 68 | 0 | 473 | 0.863 |

2) The implementation and intensity of business closure, by country.The total column shows the proportion of regions that did issue mandatory business closure orders

| Country | 0 | 0.16 | 0.33 | 0.66 | 1 | Total |
| --- | --- | --- | --- | --- | --- | --- |
| England | 299 | 0 | 0 | 0 | 0 | 0 |
| France | 127 | 0 | 0 | 0 | 143 | 1 |
| Germany | 608 | 0 | 0 | 0 | 0 | 0 |
| Iran | 548 | 0 | 0 | 0 | 0 | 0 |
| Italy | 25 | 0 | 51 | 232 | 331 | 1 |
| Netherlands | 296 | 0 | 0 | 0 | 0 | 0 |
| South Korea | 595 | 0 | 0 | 0 | 0 | 0 |
| Spain | 328 | 0 | 0 | 0 | 0 | 0 |
| Sweden | 503 | 0 | 0 | 0 | 0 | 0 |
| USA | 273 | 13 | 178 | 138 | 636 | 0.98 |

3) Summary statistics of variables

| Variable | Min | Max | Mean | sd |
| --- | --- | --- | --- | --- |
| days | 1 | 67 | 15.698 | 10.385 |
| log(days) | 0 | 4.205 | 2.47 | 0.857 |
| z(log(days)) | -2.882 | 2.024 | 0 | 1 |
| home | 0 | 29 | 4.843 | 6.831 |
| log(home+1) | 0 | 3.401 | 1.075 | 1.18 |
| z(log(home+1)) | -0.911 | 1.972 | 0 | 1 |
| busi | 0 | 32.75 | 2.874 | 5.557 |
| log(busi+1) | 0 | 3.519 | 0.657 | 1.048 |
| z(log(busi+1)) | -0.627 | 2.731 | 0 | 1 |
| mdays_s | 2 | 45.5 | 15.698 | 5.853 |
| z(mdays_s) | -2.34 | 5.092 | 0 | 1 |
| mdays_c | 10.588 | 22.495 | 15.698 | 4.116 |
| z(mdays_c) | -1.241 | 1.652 | 0 | 1 |
| mhome_s | 0 | 14 | 4.843 | 4.391 |
| z(mhome_s) | -1.103 | 2.085 | 0 | 1 |
| mhome_c | 0 | 12.601 | 4.843 | 4.215 |
| z(mhome_c) | -1.149 | 1.84 | 0 | 1 |
| mbusi_s | 0 | 15.527 | 2.874 | 4.09 |
| z(mbusi_s) | -0.703 | 3.094 | 0 | 1 |
| mbusi_c | 0 | 10.897 | 2.874 | 3.872 |
| z(mbusi_c) | -0.742 | 2.072 | 0 | 1 |

4) VIF values among all regressors in the full model

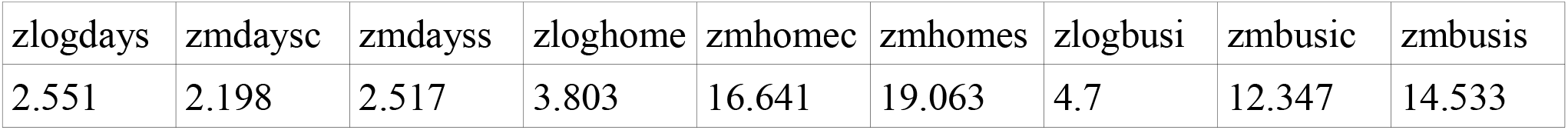

4) VIF values among all regressors in the full model.
|  |  |  |  |  |  |  |  |  |
| --- | --- | --- | --- | --- | --- | --- | --- | --- |
| zlogdays | zmdaysc | zmdayss | zloghome | zmhomec | zmhomes | zlogbusi | zmbusic | zmbusis |
| 2.551 | 2.198 | 2.517 | 3.803 | 16.641 | 19.063 | 4.7 | 12.347 | 14.533 |

5) VIF values among main regressors in the full model (cluster specific means are combinations of these)

5) VIF values among main regressors in the full model (cluster specific means are combinations of these).
|  |  |  |
| --- | --- | --- |
| zlogdays | zloghome | zlogbusi |
| 1.361 | 1.416 | 1.135 |

6) Range of estimates from the full model

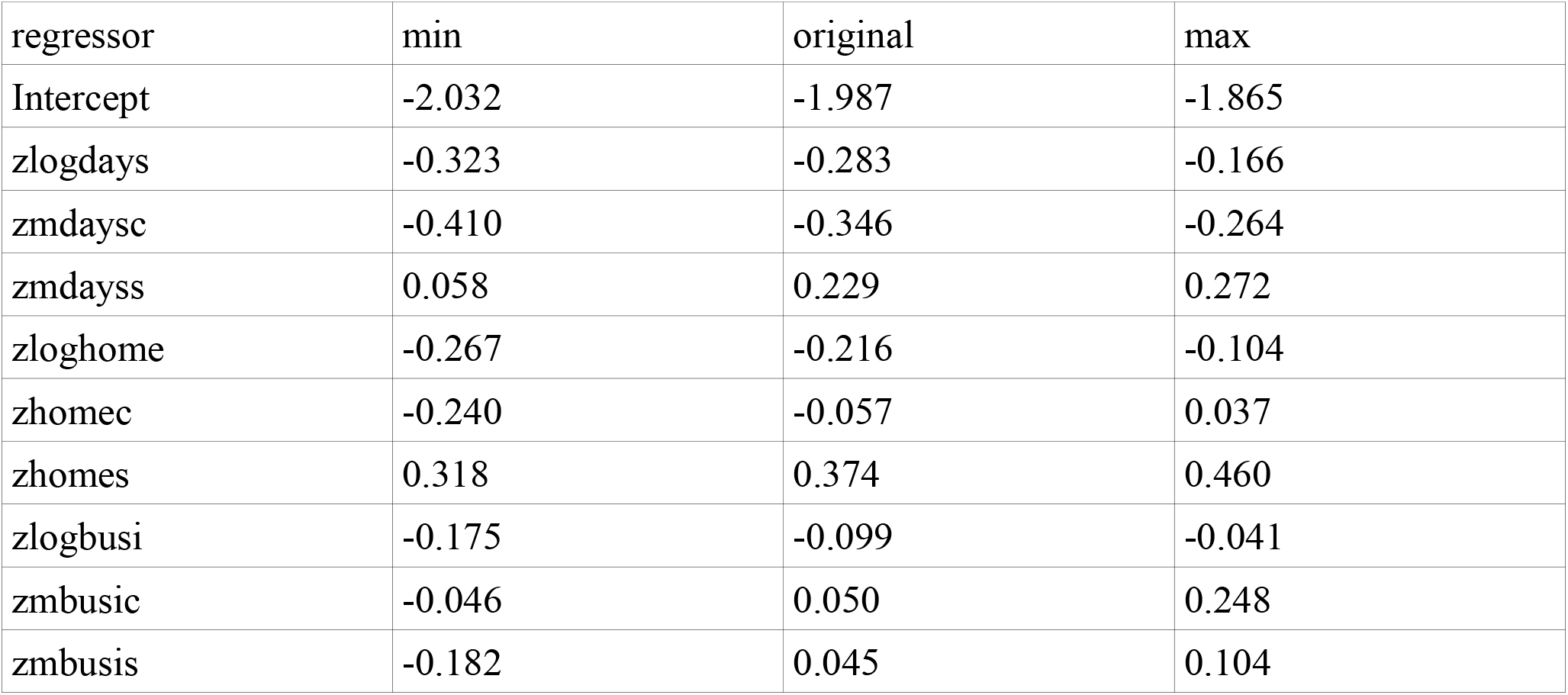

7) Distributions of random terms from the full model

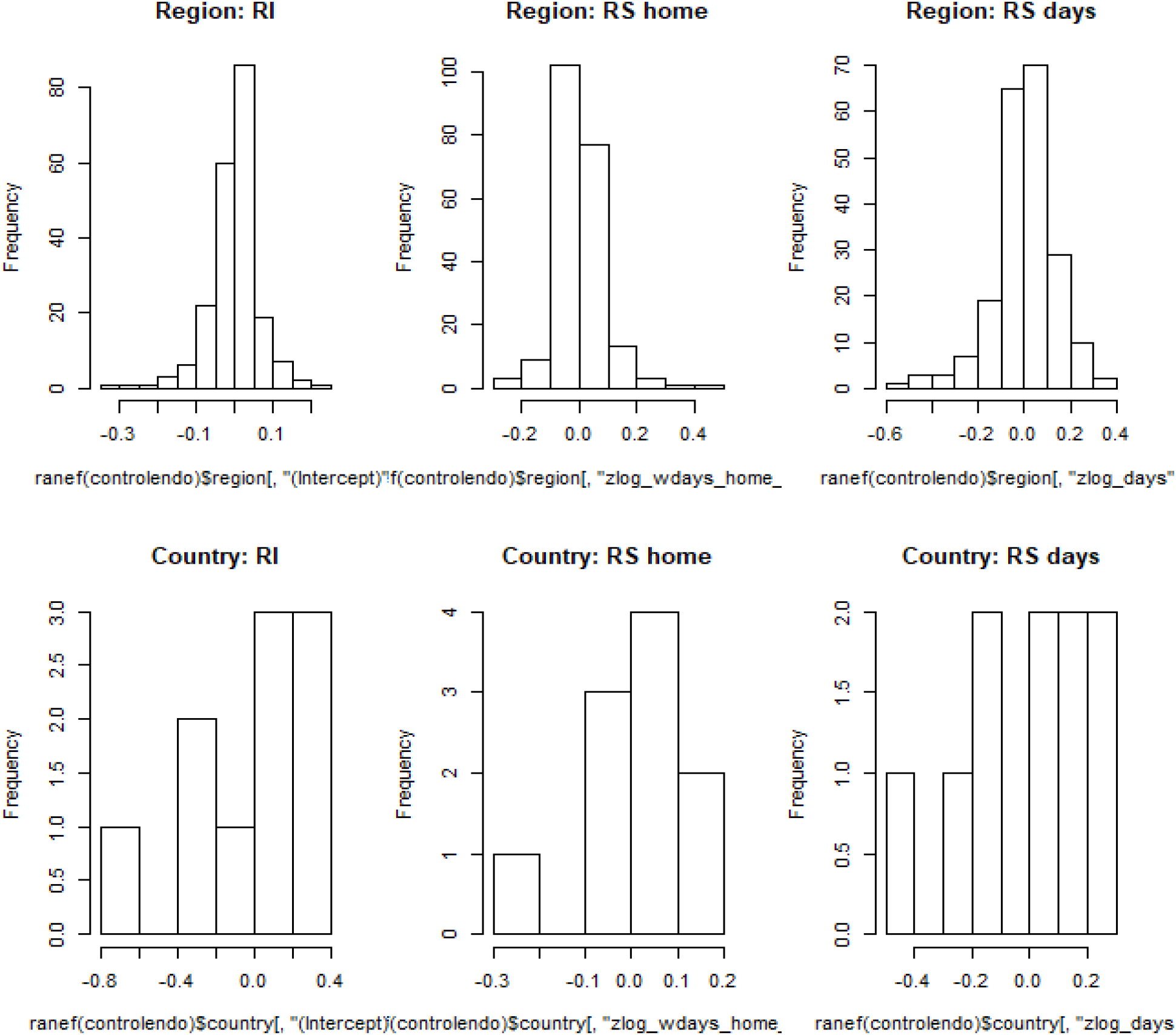

8) Dispersion parameter and oversidpersion test of the full model

| Chisq. | Df | P | Dispersion parameter |
| --- | --- | --- | --- |
| 2836.682 | 5300 | 1 | 0.535 |

9) Result of the estimation of the full model from equation 1. Correlations among random effects were estimated but are not shown for purpose of clarity

| Random effects |  |  |  |
| --- | --- | --- | --- |
| Group | Name | Std. Dev. | P-val. |
| Sub-national unit | RI | 0.090 | Not applicable |
|  | RS: days | 0.162 | <0.001 |
|  | RS: home | 0.102 | <0.001 |
| Country | RI | 0.234 | Not applicable |
|  | RS: days | 0.157 | <0.001 |
|  | RS: home | 0.106 | 0.020 |
| Number obs: 5324, Number countries: 10, Number sub-national units: 209 |  |  |  |
| Fixed effects |  |  |  |
| Name | Estimate | Std. Err. | P-val. |
| Intercept | -1.987 | 0.077 | Not applicable |
| days* | -0.283 | 0.055 | 0.002 |
| home# | -0.216 | 0.048 | 0.026 |
| busi# | -0.100 | 0.046 | 0.148 |
| mdaysc\|\| | -0.346 | 0.037 | <0.001 |
| mdayss\|\| | 0.228 | 0.026 | <0.001 |
| mhomec\|\| | -0.057 | 0.062 | 0.539 |
| mhomes\|\| | 0.374 | 0.057 | <0.001 |
| mbusic\|\| | 0.050 | 0.050 | 0.488 |
| mbusis\|\| | 0.045 | 0.052 | 0.453 |

|| standardized; \* log transformed and standardized; # log transformed after addition of one and standardized

