## Supplementary material for "Lockdown benefit varies among countries and sub-national units: a reanalysis of the data by Bendavid et al. (2021)": Functions and script for the analysis: overdisp.R

```
overdisp.test<-function(x){
  ##last updated Nov 26 2019
  ##additions/changes: bugfix for zeroinfl; deals with glmmTMB (beta and nbinom family)
  ##function needed for Gamma family:
  gamma.par.transf<-function(mean=NULL, var=NULL, shape=NULL, scale=NULL){
    ##parameterizations are
    ##shape and scale
    ##shape and mean
    ##var and mean
    if(is.null(mean) & is.null(var) & !is.null(shape) & !is.null(scale)){
      mean=shape*scale
      var=shape*scale^2
    }else if(is.null(shape) & is.null(scale) & !is.null(mean) & !is.null(var)){
      scale=var/mean
      shape=mean/scale
    }else if(is.null(scale) & is.null(var) & !is.null(shape) & !is.null(mean)){
      scale=mean/shape
      var=shape*scale^2
    }else{
      Stop("unsupported parameter commbination")
    }
    return(list(shape=as.vector(shape), scale=as.vector(scale),  mean=as.vector(mean), var=as.vector(var)))
  }
  
  if(class(x)[[1]]!="glmmTMB"){
    pr=residuals(x, type ="pearson")
    sum.dp=sum(pr^2)
  }
  n.model.terms=NULL
  if(class(x)[[1]]=="glmerMod"){
    n.model.terms=length(fixef(x))+nrow(as.data.frame(summary(x)$varcor))
    if(grepl(x=as.character(x@call)[4], pattern="negative.binomial") | grepl(x=as.character(x@call)[4], pattern="Gamma")){n.model.terms=n.model.terms+1}
  }else if(class(x)[[1]]=="glm" | class(x)[[1]]=="negbin"){
    n.model.terms=length(x$coefficients)
    n.model.terms=n.model.terms+length(summary(x)$theta)
  }else if(class(x)[[1]]=="zeroinfl"){
    n.model.terms=sum(unlist(lapply(x$coefficients, length)))
    n.model.terms=n.model.terms+length(summary(x)$theta)
  }else if(class(x)[[1]]=="betareg"){
    n.model.terms=sum(unlist(lapply(x$coefficients, length)))
  }else if(class(x)[[1]]=="glmmTMB"){
    print("caution: support for models of class glmmTMB is experimental and embryonic")
    print("and don't worry if it takes some time")
    #browser()
    #n.model.terms=sum(unlist(lapply(fixef(x), length)))+nrow(extract.ranef.from.glmmTB(x))
    n.model.terms=sum(unlist(lapply(fixef(x), length)))##fixed effects
    n.model.terms=n.model.terms+sum(unlist(lapply(summary(x)$varcor, function(y){
      if(!is.null(y)){
        lapply(y, function(yy){
          xx=dim(attr(yy, "correlation"))
          return((xx[1]^2-xx[1])/2+length(attr(yy, "stddev")))
        })
      }else{
        0
      }
    })))
    if((grepl(x=x$modelInfo$family$family, pattern="nbinom") | x$modelInfo$family$family=="beta") &
       length(attr(terms(x$call$dispformula), "term.labels"))==0){
      #n.model.terms=n.model.terms+1
      ##but maybe it needs to be done for negbin??? need to check this
    }
    pr=residuals(x, type ="response")
    if(x$modelInfo$family$family=="poisson"){
      fitted.var = fitted(x)
    }else{
      #model.terms=attr(terms(x$call$dispformula), "term.labels")#attr(terms(x$call$dispformula), "intercept")
      sigma.f=exp(as.vector(model.matrix(object=x$call$dispformula, data=x$frame)[, names(fixef(x)$disp), drop=F]%*%fixef(x)$disp))
      if(x$modelInfo$family$family=="nbinom2"){
        fitted.var = fitted(x)*(1+fitted(x)/sigma.f)#fitted(x) + fitted(x)^2/sigma(x)
      }else if(x$modelInfo$family$family=="nbinom1"){
        fitted.var = fitted(x)*(1+sigma.f)#fitted(x) + fitted(x)/sigma(x)
      }else if(x$modelInfo$family$family=="beta"){
        xfitted=fitted(x)
        shape1 = xfitted * sigma.f
        shape2 = (1 - xfitted) * sigma.f
        #xfitted(1-xfitted)/(1+sigma(x))#according to the glmmTMB help page for sigma
        fitted.var = shape1*shape2/((shape1+shape2)^2*(shape1+shape2+1))
      }else if(x$modelInfo$family$family=="Gamma"){
        #x.disp=exp(as.vector(model.matrix(object=x$call$dispformula, data=x$frame)%*%log(sigma(x))))
        xx=gamma.par.transf(mean=fitted(x), var=NULL, shape=1/sigma.f, scale=NULL)
        fitted.var = xx$var
        #xdf=length(residuals(x))-(length(coef(x))+1)
        #n.model.terms=sum(unlist(lapply(fixef(x), length)))+sum(unlist(lapply(summary(x)$varcor, function(y){length(as.data.frame(y))})))
        xdf=length(residuals(x))-n.model.terms
      }
    }
    #n.model.terms=sum(unlist(lapply(fixef(x), length)))+sum(unlist(lapply(summary(x)$varcor, function(y){length(as.data.frame(y))})))
    #xdf=length(pr)-n.model.terms
    sum.dp=sum((pr/sqrt(fitted.var))^2)
    #if(as.numeric(substr(sessionInfo()[["otherPkgs"]][["glmmTMB"]]$Version, start=1, stop=1))>0){
    #sum.dp=sum(residuals(x, type ="pearson")^2)
    #}
  }
  if(!is.null(n.model.terms)){
    xdf=length(pr)-n.model.terms
    return(data.frame(chisq=sum.dp, df=xdf, P=1-pchisq(sum.dp, xdf), dispersion.parameter=sum.dp/xdf))
  }else{
    print("model isn't of any of the currently supported classes (glm, negbin, zeroinfl, glmerMod, glmmTMB)")
  }
}
```
