## Supplementary material for "Lockdown benefit varies among countries and sub-national units: a reanalysis of the data by Bendavid et al. (2021)": Functions and script for the analysis: reanalysis_BendavidEtAl2021.R

```
### The data can be downloaded at: https://onlinelibrary.wiley.com/doi/10.1111/eci.13484
# Use one folder for all data and the R-functions. 
# Enter the path to the folder with the data and the functions 
# between the two quotation marks. Make sure to use the following slash: /
folder=""


### load data Spain
spain=read.csv(paste(folder, "/eci13484-sup-0002-supinfo/covid_npi_analysis/output/processed/ESP_processed.csv", sep="")) 

# generate cumulated cases
bla=vector()
region=as.character(unique(spain$CCAA))
for(i in 1:length(region)){
  check=spain[spain$CCAA==region[i], ]  
  bla=c(bla, cumsum(check$total))
}

# date
spain$date = as.Date(spain$date)

# remove administrative district "Total"
spain=spain[as.character(spain$adm1_name)!="Total", ]

# order according to district and ascending date
spain=spain[order(spain$adm1_name, spain$date),]

# add country name column
spain$country="Spain"

# generate id
spain$id=paste(as.character(spain$country), as.character(spain$adm1_name), as.character(spain$date))

# set business closure to 0
spain$business_closure=0

# dependent variable
# keep rows with more than 9 cumulated cases
spain=spain[spain$cum_confirmed_cases>9, ]

# logarithm of cum_confirmend_cases
spain$log_cum_confirmed_cases=log(spain$cum_confirmed_cases)

# generate difference in log_cum_confirmed_cases
bla=vector()
region=as.character(unique(spain$adm1_name))
for(i in 1:length(region)){
  check=spain[spain$adm1_name==region[i], ]  
  check=check[!is.na(check$adm1_name), ]
  bla=c(bla, c(NA, diff(check$log_cum_confirmed_cases))) 
}

spain$diff_log_cum_confirmed_cases=bla

spain_data=data.frame(cum_confirmed_cases=as.numeric(spain$cum_confirmed_cases),
                      log_cum_confirmed_cases=as.numeric(spain$log_cum_confirmed_cases),
                      diff_log_cum_confirmed_cases=as.numeric(spain$diff_log_cum_confirmed_cases),
                      date=as.Date(spain$date), 
                      adm_name=as.factor(spain$adm1_name),
                      country=as.factor(spain$country), 
                      id=as.character(spain$id), 
                      business_closure=as.numeric(spain$business_closure), 
                      home_isolation=as.numeric(spain$home_isolation))

### load data Germany
germany=read.csv(paste(folder, "/eci13484-sup-0002-supinfo/covid_npi_analysis/output/processed/GER_processed.csv", sep="")) 

# date
germany$date = as.Date(germany$date)

# order according to district and ascending date
germany=germany[order(germany$adm1_name, germany$date),]

# country
germany$country="Germany"

# generate id
germany$id=paste(as.character(germany$country), as.character(germany$adm1_name), 
                 as.character(germany$date))

# set business_closure to 0
germany$business_closure=0

# Keep rows with more than 9 cumulated cases
germany=germany[germany$cum_confirmed_cases>9, ]

# build log
germany$log_cum_confirmed_cases=log(germany$cum_confirmed_cases)

# generate difference in log_cum_confirmed_cases
bla=vector()
region=as.character(unique(germany$adm1_name))
for(i in 1:length(region)){
  check=germany[germany$adm1_name==region[i], ]  
  check=check[!is.na(check$adm1_name), ]
  bla=c(bla, c(NA, diff(check$log_cum_confirmed_cases))) 
}

germany$diff_log_cum_confirmed_cases=bla

germany_data=data.frame(cum_confirmed_cases=as.numeric(germany$cum_confirmed_cases),
                        log_cum_confirmed_cases=as.numeric(germany$log_cum_confirmed_cases),  
                        diff_log_cum_confirmed_cases=as.numeric(germany$diff_log_cum_confirmed_cases),  
                        date=as.Date(germany$date), 
                        adm_name=as.factor(germany$adm1_name),
                        country=as.factor(germany$country), 
                        id=as.character(germany$id), 
                        business_closure=as.numeric(germany$business_closure), 
                        home_isolation=as.numeric(germany$home_isolation))

### load data France
france=read.csv(paste(folder, "/eci13484-sup-0002-supinfo/covid_npi_analysis/data/processing/FRA_processed.csv", sep="")) 

# date
france$date = as.Date(france$date)

# order according to district and ascending date
france=france[order(france$adm1_name, france$date),]

# country
france$country="France"

# generate id
france$id=paste(as.character(france$country), as.character(france$adm1_name), 
                 as.character(france$date))

# Dependent variable. Work with imputed cases
# Keep rows with more than 9 cumulated cases
france=france[france$cum_confirmed_cases_imputed>9, ]
france$log_cum_confirmed_cases=log(france$cum_confirmed_cases_imputed)
france=france[!is.na(france$adm1_id), ]

# generate difference in log_cum_confirmed_cases
bla=vector()
region=levels(france$adm1_name)
for(i in 1:length(region)){
  check=france[france$adm1_name==region[i], ]  
  check=check[!is.na(check$adm1_name), ]
  bla=c(bla, c(NA, diff(check$log_cum_confirmed_cases))) 
}

france$diff_log_cum_confirmed_cases=bla

france_data=data.frame(cum_confirmed_cases=as.numeric(france$cum_confirmed_cases),
                       log_cum_confirmed_cases=as.numeric(france$log_cum_confirmed_cases),
                       diff_log_cum_confirmed_cases=as.numeric(france$diff_log_cum_confirmed_cases),
                       date=as.Date(france$date), 
                       adm_name=as.factor(france$adm1_name),
                       country=as.factor(france$country), 
                       id=as.character(france$id), 
                       business_closure=as.numeric(france$business_closure), 
                       home_isolation=as.numeric(france$home_isolation))

### load data Iran
iran=read.csv(paste(folder, "/eci13484-sup-0002-supinfo/covid_npi_analysis/data/processing/IRN_processed.csv", sep="")) 

# date
iran$date = as.Date(iran$date)

# order according to district and ascending date
iran=iran[order(iran$adm1_name, iran$date),]

# country
iran$country="Iran"

# generate id
iran$id=paste(as.character(iran$country), as.character(iran$adm1_name), 
                as.character(iran$date))

# set business_closure to 0
iran$business_closure=0 

# Dependent variable. Work with imputed cases
# Keep rows with more than 9 cumulated cases
iran=iran[iran$cum_confirmed_cases_imputed>9, ]
iran$log_cum_confirmed_cases=log(iran$cum_confirmed_cases_imputed)

# generate difference in log_cum_confirmed_cases
bla=vector()
region=levels(iran$adm1_name)
for(i in 1:length(region)){
  check=iran[iran$adm1_name==region[i], ]  
  check=check[!is.na(check$adm1_name), ]
  bla=c(bla, c(NA, diff(check$log_cum_confirmed_cases))) 
}

iran$diff_log_cum_confirmed_cases=bla

iran_data=data.frame(cum_confirmed_cases=as.numeric(iran$cum_confirmed_cases),
                     log_cum_confirmed_cases=as.numeric(iran$log_cum_confirmed_cases),
                     diff_log_cum_confirmed_cases=as.numeric(iran$diff_log_cum_confirmed_cases),
                     date=as.Date(iran$date), 
                     adm_name=as.factor(iran$adm1_name),
                     country=as.factor(iran$country), 
                     id=as.character(iran$id), 
                     business_closure=as.numeric(iran$business_closure), 
                     home_isolation=as.numeric(iran$home_isolation))

### load data Italy
italy=read.csv(paste(folder, "/eci13484-sup-0002-supinfo/covid_npi_analysis/data/processing/ITA_processed.csv", sep="")) 

# date
italy$date = as.Date(italy$date)

# country
italy$country="Italy"

# order according to district and ascending date
italy=italy[order(italy$adm1_name, italy$date),]

# generate id
italy$id=paste(as.character(italy$country), as.character(italy$adm1_name), 
              as.character(italy$date))
sum(duplicated(italy$id))

# Keep maximal cum_case for duplicated
italy=italy[order(italy$id, -abs(italy$cum_confirmed_cases_imputed) ), ]
italy=italy[!duplicated(italy$id), ]

# order according to district and ascending date
italy=italy[order(italy$adm1_name, italy$date),]

# Dependent variable. Work with imputed cases
# Keep rows with more than 9 cumulated cases
italy=italy[italy$cum_confirmed_cases_imputed>9, ]
italy$log_cum_confirmed_cases=log(italy$cum_confirmed_cases_imputed)

# generate difference in log_cum_confirmed_cases
bla=vector()
region=levels(italy$adm1_name)
for(i in 1:length(region)){
  check=italy[italy$adm1_name==region[i], ]  
  check=check[!is.na(check$adm1_name), ]
  bla=c(bla, c(NA, diff(check$log_cum_confirmed_cases))) 
}

italy$diff_log_cum_confirmed_cases=bla

italy_data=data.frame(cum_confirmed_cases=as.numeric(italy$cum_confirmed_cases),
                      log_cum_confirmed_cases=as.numeric(italy$log_cum_confirmed_cases),
                      diff_log_cum_confirmed_cases=as.numeric(italy$diff_log_cum_confirmed_cases),
                      date=as.Date(italy$date), 
                      adm_name=as.factor(italy$adm1_name),
                      country=as.factor(italy$country), 
                      id=as.character(italy$id), 
                      business_closure=as.numeric(italy$business_closure), 
                      home_isolation=as.numeric(italy$home_isolation))

### load data South Korea
corea=read.csv(paste(folder, "/eci13484-sup-0002-supinfo/covid_npi_analysis/data/processing/KOR_processed.csv", sep="")) 

# date
corea$date = as.Date(corea$date)

# country
corea$country="Corea"

# order according to district and ascending date
corea=corea[order(corea$adm1_name, corea$date),]

# generate id
corea$id=paste(as.character(corea$country), as.character(corea$adm1_name), 
               as.character(corea$date))

# set home_isolation to 0
corea$home_isolation=0

# set business closure to 0
corea$business_closure=0

# Dependent variable. 
# Keep rows with more than 9 cumulated cases
corea=corea[corea$cum_confirmed_cases>9, ]
corea$log_cum_confirmed_cases=log(corea$cum_confirmed_cases)

# generate difference in log_cum_confirmed_cases
bla=vector()
region=levels(corea$adm1_name)
for(i in 1:length(region)){
  check=corea[corea$adm1_name==region[i], ]  
  check=check[!is.na(check$adm1_name), ]
  bla=c(bla, c(NA, diff(check$log_cum_confirmed_cases))) 
}

corea$diff_log_cum_confirmed_cases=bla

corea_data=data.frame(cum_confirmed_cases=as.numeric(corea$cum_confirmed_cases),
                      log_cum_confirmed_cases=as.numeric(corea$log_cum_confirmed_cases),
                      diff_log_cum_confirmed_cases=as.numeric(corea$diff_log_cum_confirmed_cases),
                      date=as.Date(corea$date), 
                      adm_name=as.factor(corea$adm1_name),
                      id=as.character(corea$id), 
                      country=as.factor(corea$country), 
                      business_closure=as.numeric(corea$business_closure), 
                      home_isolation=as.numeric(corea$home_isolation))

### load data Netherland
netherland=read.csv(paste(folder, "/eci13484-sup-0002-supinfo/covid_npi_analysis/output/processed/NL_processed.csv", sep="")) 

# date
netherland$date = as.Date(netherland$date)

# remove administrative district without name
netherland=netherland[!is.na(netherland$adm1_name), ]

# country
netherland$country="Netherland"

# order according to district and ascending date
netherland=netherland[order(netherland$adm1_name, netherland$date),]

# generate id
netherland$id=paste(as.character(netherland$country), as.character(netherland$adm1_name), 
               as.character(netherland$date))

# set business_closure to 0
netherland$business_closure=0

# Dependent variable. 
# Keep rows with more than 9 cumulated cases
netherland=netherland[netherland$cum_confirmed_cases>9, ]
netherland$log_cum_confirmed_cases=log(netherland$cum_confirmed_cases)

# generate difference in log_cum_confirmed_cases
bla=vector()
region=levels(netherland$adm1_name)
for(i in 1:length(region)){
  check=netherland[netherland$adm1_name==region[i], ]  
  check=check[!is.na(check$adm1_name), ]
  bla=c(bla, c(NA, diff(check$log_cum_confirmed_cases))) 
}

netherland$diff_log_cum_confirmed_cases=bla

netherland_data=data.frame(cum_confirmed_cases=as.numeric(netherland$cum_confirmed_cases),
                      log_cum_confirmed_cases=as.numeric(netherland$log_cum_confirmed_cases),
                      diff_log_cum_confirmed_cases=as.numeric(netherland$diff_log_cum_confirmed_cases),
                      date=as.Date(netherland$date), 
                      adm_name=as.factor(netherland$adm1_name),
                      country=as.factor(netherland$country), 
                      id=as.character(netherland$id), 
                      business_closure=as.numeric(netherland$business_closure), 
                      home_isolation=as.numeric(netherland$home_isolation))

### load data Sweden
sweden=read.csv(paste(folder, "/eci13484-sup-0002-supinfo/covid_npi_analysis/output/processed/SWE_processed.csv", sep="")) 

# date
sweden$date = as.Date(sweden$date)

# country
sweden$country="Sweden"

# order according to district and ascending date
sweden=sweden[order(sweden$adm1_name, sweden$date),]

# generate id
sweden$id=paste(as.character(sweden$country), as.character(sweden$adm1_name), 
                    as.character(sweden$date))

# set home_isolation to 0
sweden$home_isolation=0

# set business_closure to 0
sweden$business_closure=0

# Dependent variable. 
# Keep rows with more than 9 cumulated cases
sweden=sweden[sweden$cum_confirmed_cases>9, ]
sweden$log_cum_confirmed_cases=log(sweden$cum_confirmed_cases)

# generate difference in log_cum_confirmed_cases
bla=vector()
region=levels(sweden$adm1_name)
for(i in 1:length(region)){
  check=sweden[sweden$adm1_name==region[i], ]  
  check=check[!is.na(check$adm1_name), ]
  bla=c(bla, c(NA, diff(check$log_cum_confirmed_cases))) 
}

sweden$diff_log_cum_confirmed_cases=bla

sweden_data=data.frame(cum_confirmed_cases=as.numeric(sweden$cum_confirmed_cases),
                       log_cum_confirmed_cases=as.numeric(sweden$log_cum_confirmed_cases),
                       diff_log_cum_confirmed_cases=as.numeric(sweden$diff_log_cum_confirmed_cases),
                       date=as.Date(sweden$date), 
                       adm_name=as.factor(sweden$adm1_name),
                       country=as.factor(sweden$country), 
                       id=as.character(sweden$id), 
                       business_closure=as.numeric(sweden$business_closure), 
                       home_isolation=as.numeric(sweden$home_isolation))

### load data UK
uk=read.csv(paste(folder, "/eci13484-sup-0002-supinfo/covid_npi_analysis/output/processed/UK_processed.csv", sep="")) 

# date
uk$date = as.Date(uk$date)

# reorder data based on increasing date (actually based on decreasing date)
uk=uk[order(uk$adm1_name, uk$date),]

# country
uk$country="UK"

# id
uk$id=paste(as.character(uk$country), as.character(uk$adm1_name), 
                as.character(uk$date))

# set business_closure to 0
uk$business_closure=0

# Dependent variable. 
# Keep rows with more than 9 cumulated cases
uk=uk[uk$cum_confirmed_cases>9, ]
uk$log_cum_confirmed_cases=log(uk$cum_confirmed_cases)

# generate difference in log_cum_confirmed_cases
bla=vector()
region=levels(uk$adm1_name)
for(i in 1:length(region)){
  check=uk[uk$adm1_name==region[i], ]  
  check=check[!is.na(check$adm1_name), ]
  bla=c(bla, c(NA, diff(check$log_cum_confirmed_cases))) 
}

uk$diff_log_cum_confirmed_cases=bla

uk_data=data.frame(cum_confirmed_cases=as.numeric(uk$cum_confirmed_cases),
                   log_cum_confirmed_cases=as.numeric(uk$log_cum_confirmed_cases),
                   diff_log_cum_confirmed_cases=as.numeric(uk$diff_log_cum_confirmed_cases),
                   date=as.Date(uk$date), 
                   adm_name=as.factor(uk$adm1_name),
                   country=as.factor(uk$country), 
                   id=as.character(uk$id), 
                   business_closure=as.numeric(uk$business_closure), 
                   home_isolation=as.numeric(uk$home_isolation))

### load data USA
usa=read.csv(paste(folder, "/eci13484-sup-0002-supinfo/covid_npi_analysis/data/processing/USA_processed.csv", sep="")) 

# date
usa$date = as.Date(usa$date)

# country
usa$country="USA"

# reorder data based on increasing date (actually based on decreasing date)
usa=usa[order(usa$adm1_name, usa$date),]

# id
usa$id=paste(as.character(usa$country), as.character(usa$adm1_name), 
            as.character(usa$date))

# Dependent variable. Work with imputed cumulated cases
# Keep rows with more than 9 cumulated cases
usa=usa[usa$cum_confirmed_cases_imputed>9, ]
usa$log_cum_confirmed_cases=log(usa$cum_confirmed_cases_imputed)

# generate difference in log_cum_confirmed_cases
bla=vector()
region=levels(usa$adm1_name)
for(i in 1:length(region)){
  check=usa[usa$adm1_name==region[i], ]  
  check=check[!is.na(check$adm1_name), ]
  bla=c(bla, c(NA, diff(check$log_cum_confirmed_cases))) 
}

usa$diff_log_cum_confirmed_cases=bla

usa_data=data.frame(cum_confirmed_cases=as.numeric(usa$cum_confirmed_cases),
                    log_cum_confirmed_cases=as.numeric(usa$log_cum_confirmed_cases),
                    diff_log_cum_confirmed_cases=as.numeric(usa$diff_log_cum_confirmed_cases),
                    date=as.Date(usa$date), 
                    adm_name=as.factor(usa$adm1_name),
                    country=as.factor(usa$country), 
                    id=as.factor(usa$id), 
                    business_closure=as.numeric(usa$business_closure), 
                    home_isolation=as.numeric(usa$home_isolation))

### Combine data for further analysis
ana_data=rbind(spain_data, germany_data, france_data, corea_data, iran_data,
               italy_data, netherland_data, sweden_data, uk_data,
               usa_data)

# remove missing values
ana_data2=na.omit(ana_data)

# remove cases based on values in dependent variable that are not possible
ana_data3=ana_data2[ana_data2$diff_log_cum_confirmed_cases>=0, ]

### generate region id
ana_data3$region=paste(ana_data3$country, ana_data3$adm_name)


### generate days of measure implementation: home_isolation
home_isolation_region=vector()
home_isolation_date=as.Date(vector())
home_isolation_days=vector()
home_isolation_wdays=vector()

region=unique(as.character(ana_data3$region))
for(i in 1:length(region)){
  check=ana_data3[as.character(ana_data3$region)==region[i], ]   
  checkb=check[check$home_isolation>0, ]  
  if(nrow(checkb)>0){
    home_isolation_region=c(home_isolation_region, rep(region[i], length.out=nrow(checkb)))
    home_isolation_start=checkb$date[1]
    home_isolation_date=c(home_isolation_date, as.Date(checkb$date))
    home_isolation_days=c(home_isolation_days, 
                           cumsum(checkb$home_isolation>0))
    home_isolation_wdays=c(home_isolation_wdays, 
                           cumsum(checkb$home_isolation))
    }else{
  }
}

home_isolation_id=paste(as.character(home_isolation_region), 
                             as.character(home_isolation_date))

ana_data3$days_home_isolation=0
ana_data3$days_home_isolation[ana_data3$id %in% home_isolation_id]=home_isolation_days
ana_data3$wdays_home_isolation=0
ana_data3$wdays_home_isolation[ana_data3$id %in% home_isolation_id]=home_isolation_wdays

# check if correct
sum(is.na(ana_data3))
edit(cbind(as.character(ana_data3$adm_name), as.character(ana_data3$date), 
           ana_data3$home_isolation, ana_data3$days_home_isolation, ana_data3$wdays_home_isolation))


### Generate days of measure implementation: business_closure
business_closure_region=vector()
business_closure_date=as.Date(vector())
business_closure_days=vector()
business_closure_wdays=vector()

region=unique(as.character(ana_data3$region))
for(i in 1:length(region)){
  check=ana_data3[as.character(ana_data3$region)==region[i], ]  
  checkb=check[check$business_closure>0, ]  
  if(nrow(checkb)>0){
    business_closure_region=c(business_closure_region, rep(region[i], length.out=nrow(checkb)))
    business_closure_start=checkb$date[1]
    business_closure_date=c(business_closure_date, as.Date(checkb$date))
    business_closure_days=c(business_closure_days, 
                            cumsum(checkb$business_closure>0))
    business_closure_wdays=c(business_closure_wdays, 
                            cumsum(checkb$business_closure))
    }else{
  }
}

business_closure_id=paste(as.character(business_closure_region), 
                        as.character(business_closure_date))

ana_data3$days_business_closure=0
ana_data3$days_business_closure[ana_data3$id %in% business_closure_id]=business_closure_days
ana_data3$wdays_business_closure=0
ana_data3$wdays_business_closure[ana_data3$id %in% business_closure_id]=business_closure_wdays

# check if correct
sum(is.na(ana_data3))
edit(cbind(as.character(ana_data3$adm_name), as.character(ana_data3$date), 
           ana_data3$business_closure, ana_data3$days_business_closure, ana_data3$wdays_business_closure))


### Generate epidemic age (timetrend)
days_region=vector()
days_date=as.Date(vector())
days_days=vector()

region=unique(as.character(ana_data3$region))
for(i in 1:length(region)){
  check=ana_data3[as.character(ana_data3$region)==region[i], ]  
    days_region=c(days_region, rep(region[i], length.out=nrow(check)))
    days_date=c(days_date, as.Date(check$date))
    days_days=c(days_days, c(as.numeric(check$date)-(min(as.numeric(check$date)-1))))
}

days_id=paste(as.character(days_region), 
                          as.character(days_date))

ana_data3$days=0
ana_data3$days[ana_data3$id %in% days_id]=days_days

# check if correct
sum(is.na(ana_data3))
edit(cbind(as.character(ana_data3$adm_name), as.character(ana_data3$date), 
           ana_data3$days, ana_data3$days_home_isolation, ana_data3$home_isolation))


### Get cases used for comparative analysis in Bendavid et al. (2021)
# load data Spain
spain_ori=read.csv(paste(folder, "/eci13484-sup-0002-supinfo/covid_npi_analysis/output/regression/ESP_reg_data_10.csv", sep="")) 
spain_ori$country="Spain"
spain_ori=spain_ori[! spain_ori$adm1_name=="Total", ]
spain_ori$id=paste(spain_ori$country, spain_ori$adm1_name, spain_ori$date)

# load data France
france_ori=read.csv(paste(folder, "/eci13484-sup-0002-supinfo/covid_npi_analysis/output/regression/FRA_reg_data_10.csv", sep="")) 
france_ori$country="France"
france_ori$id=paste(france_ori$country, france_ori$adm1_name, france_ori$date)

# load data Germany
germany_ori=read.csv(paste(folder, "/eci13484-sup-0002-supinfo/covid_npi_analysis/output/regression/GER_reg_data_10.csv", sep="")) 
germany_ori$country="Germany"
germany_ori$id=paste(germany_ori$country, germany_ori$adm1_name, germany_ori$date)

# load data Iran
iran_ori=read.csv(paste(folder, "/eci13484-sup-0002-supinfo/covid_npi_analysis/output/regression/IRN_reg_data_10.csv", sep="")) 
iran_ori$country="Iran"
iran_ori$id=paste(iran_ori$country, iran_ori$adm1_name, iran_ori$date)

# load data Italy
italy_ori=read.csv(paste(folder, "/eci13484-sup-0002-supinfo/covid_npi_analysis/output/regression/ITA_reg_data_10.csv", sep="")) 
italy_ori$country="Italy"
italy_ori=italy_ori[order(italy_ori$adm1_name, italy_ori$date),]
# generate id
italy_ori$id=paste(as.character(italy_ori$country), as.character(italy_ori$adm1_name), 
               as.character(italy_ori$date))
sum(duplicated(italy_ori$id))
# Keep maximal cum_case for duplicated
italy_ori=italy_ori[order(italy_ori$id, -abs(italy_ori$cum_confirmed_cases_imputed) ), ]
italy_ori=italy_ori[!duplicated(italy_ori$id), ]

# load data Korea
corea_ori=read.csv(paste(folder, "/eci13484-sup-0002-supinfo/covid_npi_analysis/output/regression/KOR_reg_data_10.csv", sep="")) 
corea_ori$country="Corea"
corea_ori$id=paste(corea_ori$country, corea_ori$adm1_name, corea_ori$date)

# load data Netherland
netherland_ori=read.csv(paste(folder, "/eci13484-sup-0002-supinfo/covid_npi_analysis/output/regression/NL_reg_data_10.csv", sep="")) 
netherland_ori$country="Netherland"
netherland_ori$id=paste(netherland_ori$country, netherland_ori$adm1_name, netherland_ori$date)

# load data Sweden
sweden_ori=read.csv(paste(folder, "/eci13484-sup-0002-supinfo/covid_npi_analysis/output/regression/SWE_reg_data_10.csv", sep="")) 
sweden_ori$country="Sweden"
sweden_ori$id=paste(sweden_ori$country, sweden_ori$adm1_name, sweden_ori$date)

# load data England
uk_ori=read.csv(paste(folder, "/eci13484-sup-0002-supinfo/covid_npi_analysis/output/regression/UK_reg_data_10.csv", sep="")) 
uk_ori$country="UK"
uk_ori$id=paste(uk_ori$country, uk_ori$adm1_name, uk_ori$date)

# load data USA
usa_ori=read.csv(paste(folder, "/eci13484-sup-0002-supinfo/covid_npi_analysis/output/regression/USA_reg_data_10.csv", sep="")) 
usa_ori$country="USA"
usa_ori$id=paste(usa_ori$country, usa_ori$adm1_name, usa_ori$date)

# combine data
ori_ana_id=c(usa_ori$id, uk_ori$id, sweden_ori$id, iran_ori$id, corea_ori$id, 
             italy_ori$id, france_ori$id, germany_ori$id, netherland_ori$id, 
             spain_ori$id)

### Select cases used for comparative analysis in Bendavid et al. (2021)
ana_data4=ana_data3[ana_data3$id %in% ori_ana_id, ]


### Generate cluster means
### Mean days home isolation, region
days_region=vector()
days_date=as.Date(vector())
mean_hdays_region=vector()

region=unique(as.character(ana_data4$region))
for(i in 1:length(region)){
  check=ana_data4[as.character(ana_data4$region)==region[i], ]  
  days_region=c(days_region, rep(region[i], length.out=nrow(check)))
  days_date=c(days_date, as.Date(check$date))
  mean_hdays_region=c(mean_hdays_region, rep(mean(check$wdays_home_isolation), 
                                             length.out=nrow(check)))
}

days_id=paste(as.character(days_region), 
              as.character(days_date))

ana_data4$mean_hdays_region=0
ana_data4$mean_hdays_region[ana_data4$id %in% days_id]=mean_hdays_region

# check if correct
sum(is.na(ana_data4))
edit(cbind(as.character(ana_data4$adm_name), as.character(ana_data4$date), 
           ana_data4$days, ana_data4$days_home_isolation, ana_data4$mean_hdays_region))


### Mean days home isolation, country
days_region=vector()
days_date=as.Date(vector())
mean_hdays_country=vector()

countries=unique(as.character(ana_data4$country))
for(i in 1:length(countries)){
  check=ana_data4[as.character(ana_data4$country)==countries[i], ]  
  days_region=c(days_region, check$region)
  days_date=c(days_date, as.Date(check$date))
  mean_hdays_country=c(mean_hdays_country, rep(mean(check$wdays_home_isolation), 
                                               length.out=nrow(check)))
}

days_id=paste(as.character(days_region), 
              as.character(days_date))

ana_data4$mean_hdays_country=0
ana_data4$mean_hdays_country[ana_data4$id %in% days_id]=mean_hdays_country

# check if correct
sum(is.na(ana_data4))
edit(cbind(as.character(ana_data4$adm_name), as.character(ana_data4$date), 
           ana_data4$days, ana_data4$days_home_isolation, ana_data4$mean_hdays_country))


### Mean days business closure, region
days_region=vector()
days_date=as.Date(vector())
mean_bdays_region=vector()

region=unique(as.character(ana_data4$region))
for(i in 1:length(region)){
  check=ana_data4[as.character(ana_data4$region)==region[i], ]  
  days_region=c(days_region, rep(region[i], length.out=nrow(check)))
  days_date=c(days_date, as.Date(check$date))
  mean_bdays_region=c(mean_bdays_region, rep(mean(check$wdays_business_closure), 
                                             length.out=nrow(check)))
}

days_id=paste(as.character(days_region), 
              as.character(days_date))

ana_data4$mean_bdays_region=0
ana_data4$mean_bdays_region[ana_data4$id %in% days_id]=mean_bdays_region

# check if correct
sum(is.na(ana_data4))
edit(cbind(as.character(ana_data4$adm_name), as.character(ana_data4$date), 
           ana_data4$days, ana_data4$wdays_business_closure, ana_data4$mean_bdays_region))


### Mean days business closure, country
days_region=vector()
days_date=as.Date(vector())
mean_bdays_country=vector()

countries=unique(as.character(ana_data4$country))
for(i in 1:length(countries)){
  check=ana_data4[as.character(ana_data4$country)==countries[i], ]  
  days_region=c(days_region, check$region)
  days_date=c(days_date, as.Date(check$date))
  mean_bdays_country=c(mean_bdays_country, rep(mean(check$wdays_business_closure), 
                                               length.out=nrow(check)))
}

days_id=paste(as.character(days_region), 
              as.character(days_date))

ana_data4$mean_bdays_country=0
ana_data4$mean_bdays_country[ana_data4$id %in% days_id]=mean_bdays_country

# check if correct
sum(is.na(ana_data4))
edit(cbind(as.character(ana_data4$adm_name), as.character(ana_data4$date), 
           ana_data4$days, ana_data4$wdays_business_closure, ana_data4$mean_bdays_country))


### Mean days, region
days_region=vector()
days_date=as.Date(vector())
mdays_region=vector()

region=unique(as.character(ana_data4$region))
for(i in 1:length(region)){
  check=ana_data4[as.character(ana_data4$region)==region[i], ]  
  days_region=c(days_region, rep(region[i], length.out=nrow(check)))
  days_date=c(days_date, as.Date(check$date))
  mdays_region=c(mdays_region, rep(mean(check$days), 
                                             length.out=nrow(check)))
}

days_id=paste(as.character(days_region), 
              as.character(days_date))

ana_data4$mdays_region=0
ana_data4$mdays_region[ana_data4$id %in% days_id]=mdays_region

# check if correct
sum(is.na(ana_data4))
edit(cbind(as.character(ana_data4$adm_name), as.character(ana_data4$date), 
           ana_data4$days, ana_data4$mdays_region))


### Mean days, country
days_region=vector()
days_date=as.Date(vector())
mdays_country=vector()

for(i in 1:length(countries)){
  check=ana_data4[as.character(ana_data4$country)==countries[i], ]  
  days_region=c(days_region, check$region)
  days_date=c(days_date, as.Date(check$date))
  mdays_country=c(mdays_country, rep(mean(check$days), 
                                               length.out=nrow(check)))
}

days_id=paste(as.character(days_region), 
              as.character(days_date))

ana_data4$mdays_country=0
ana_data4$mdays_country[ana_data4$id %in% days_id]=mdays_country

# check if correct
sum(is.na(ana_data4))
edit(cbind(as.character(ana_data4$adm_name), as.character(ana_data4$date), 
           ana_data4$days, ana_data4$mdays_country))


### Transform variables
# home isolation
ana_data4$log_days_home_isolation=log(ana_data4$days_home_isolation+1)
ana_data4$log_wdays_home_isolation=log(ana_data4$wdays_home_isolation+1)
ana_data4$zlog_wdays_home_isolation=scale(ana_data4$log_wdays_home_isolation)
ana_data4$zmean_hdays_country=scale(ana_data4$mean_hdays_country)
ana_data4$zmean_hdays_region=scale(ana_data4$mean_hdays_region)

# business closure
ana_data4$log_days_business_closure=log(ana_data4$days_business_closure+1)
ana_data4$log_wdays_business_closure=log(ana_data4$wdays_business_closure+1)
ana_data4$zlog_wdays_business_closure=scale(ana_data4$log_wdays_business_closure)
ana_data4$zmean_bdays_country=scale(ana_data4$mean_bdays_country)
ana_data4$zmean_bdays_region=scale(ana_data4$mean_bdays_region)

# days
ana_data4$log_days=log(ana_data4$days)
ana_data4$zlog_days=scale(ana_data4$log_days)
ana_data4$zmdays_region=scale(ana_data4$mdays_region)
ana_data4$zmdays_country=scale(ana_data4$mdays_country)

### dependent variable is highly skewed!
hist(ana_data4$diff_log_cum_confirmed_cases)

# generate new dependent variable for gamma regression (with 0=min/10)
ana_data4$new_outcome=ana_data4$diff_log_cum_confirmed_cases
ana_data4$new_outcome[ana_data4$diff_log_cum_confirmed_cases==0]=
  min(ana_data4$diff_log_cum_confirmed_cases[ana_data4$diff_log_cum_confirmed_cases>0])/10

# check result
hist(ana_data4$new_outcome)


### Get descriptive stats
### Implementation and intensity of measures (by country)
implementation_home=table(ana_data4$country, ana_data4$home_isolation)
implementation_busi=table(ana_data4$country, ana_data4$business_closure)

# get regions that did not implement home isolation
region=unique(ana_data4$region)
yesh=rep(NA, times=length(region))

for(i in 1:length(region)){
  check=ana_data4[ana_data4$region==region[i], ]
  bla=as.numeric(names(table(check$wdays_home_isolation)))
  if(sum(bla)==0){
    yesh[i]=0
  }else{
    yesh[i]=1
  }
}

# get regions that did not implement business closure
yesb=rep(NA, times=length(region))

for(i in 1:length(region)){
  check=ana_data4[ana_data4$region==region[i], ]
  bla=as.numeric(names(table(check$wdays_business_closure)))
  if(sum(bla)==0){
    yesb[i]=0
  }else{
    yesb[i]=1
  }
}

implementation_region=data.frame(region=as.character(region), 
                                    isolation=as.numeric(yesh), 
                                    closure=as.numeric(yesb))

countries=unique(as.character(ana_data4$country))
share_imp_h=rep(NA, times=length(countries))
share_imp_b=rep(NA, times=length(countries))
for(i in 1:length(countries)){
  check=implementation_region[grep(countries[i], implementation_region$region), ]
  share_imp_h[i]=sum(check$isolation)/nrow(check)
  share_imp_b[i]=sum(check$closure)/nrow(check)
}

implementation_home=cbind(implementation_home, round(share_imp_h, 3))
colnames(implementation_home)=c("0", "0.33", "0.33000001", "0.5", "0.67", "0.67000002", "1", "Total")
write.csv(implementation_home, "C:/Arbeit/Covid/eci13484-sup-0002-supinfo/covid_npi_analysis/MyAnalysis/implementation_home.csv")

implementation_busi=cbind(implementation_busi, round(share_imp_b, 3))
colnames(implementation_busi)=c("0", "0.16", "0.33", "0.66", "0.67", "1", "Total")


### Summary stats
summarystats=
rbind(c(range(ana_data4$days), mean(ana_data4$days), sd(ana_data4$days)), 
c(range(ana_data4$log_days), mean(ana_data4$log_days), sd(ana_data4$log_days)), 
c(range(ana_data4$zlog_days), mean(ana_data4$zlog_days), sd(ana_data4$zlog_days)),
c(range(ana_data4$wdays_home_isolation), mean(ana_data4$wdays_home_isolation), 
  sd(ana_data4$wdays_home_isolation)),
c(range(ana_data4$log_wdays_home_isolation), mean(ana_data4$log_wdays_home_isolation), 
  sd(ana_data4$log_wdays_home_isolation)),
c(range(ana_data4$zlog_wdays_home_isolation), mean(ana_data4$zlog_wdays_home_isolation), 
  sd(ana_data4$zlog_wdays_home_isolation)),
c(range(ana_data4$wdays_business_closure), mean(ana_data4$wdays_business_closure), 
  sd(ana_data4$wdays_business_closure)),
c(range(ana_data4$log_wdays_business_closure), mean(ana_data4$log_wdays_business_closure), 
  sd(ana_data4$log_wdays_business_closure)),
c(range(ana_data4$zlog_wdays_business_closure), mean(ana_data4$zlog_wdays_business_closure), 
  sd(ana_data4$zlog_wdays_business_closure)),
c(range(ana_data4$mdays_region), mean(ana_data4$mdays_region), 
  sd(ana_data4$mdays_region)),
c(range(ana_data4$zmdays_region), mean(ana_data4$zmdays_region), 
  sd(ana_data4$zmdays_region)),
c(range(ana_data4$mdays_country), mean(ana_data4$mdays_country), 
  sd(ana_data4$mdays_country)),
c(range(ana_data4$zmdays_country), mean(ana_data4$zmdays_country), 
  sd(ana_data4$zmdays_country)),
c(range(ana_data4$mean_hdays_region), mean(ana_data4$mean_hdays_region), 
  sd(ana_data4$mean_hdays_region)),
c(range(ana_data4$zmean_hdays_region), mean(ana_data4$zmean_hdays_region), 
  sd(ana_data4$zmean_hdays_region)),
c(range(ana_data4$mean_hdays_country), mean(ana_data4$mean_hdays_country), 
  sd(ana_data4$mean_hdays_country)),
c(range(ana_data4$zmean_hdays_country), mean(ana_data4$zmean_hdays_country), 
  sd(ana_data4$zmean_hdays_country)),
c(range(ana_data4$mean_bdays_region), mean(ana_data4$mean_bdays_region), 
  sd(ana_data4$mean_bdays_region)),
c(range(ana_data4$zmean_bdays_region), mean(ana_data4$zmean_bdays_region), 
  sd(ana_data4$zmean_bdays_region)),
c(range(ana_data4$mean_bdays_country), mean(ana_data4$mean_bdays_country), 
  sd(ana_data4$mean_bdays_country)),
c(range(ana_data4$zmean_bdays_country), mean(ana_data4$zmean_bdays_country), 
  sd(ana_data4$zmean_bdays_country))
)
summarystats=round(summarystats, 3)

summarystats=cbind(
  c("days", "log(days)", "z(log(days))", "home", "log(home+1)", "z(log(home+1))",
    "busi", "log(busi+1)", "z(log(busi+1))", "mdays_s", "z(mdays_s)", "mdays_c",
    "z(mdays_c)", "mhome_s", "z(mhome_s)", "mhome_c", "z(mhome_c)", "mbusi_s",
    "z(mbusi_s)", "mbusi_c", "z(mbusi_c)"),
  summarystats)

colnames(summarystats)=c("Variable", "Min", "Max", "Mean", "sd")
summarystats=as.data.frame(summarystats)


### Run model
# Gamma-regression
# lme4 package needed
library(lme4)

# Gamma mixed-effects
control=glmer(new_outcome~
                    zlog_days+
                    zlog_wdays_home_isolation+
                    zlog_wdays_business_closure+
                    (1+zlog_days+zlog_wdays_home_isolation|region)+
                    (1+zlog_days+zlog_wdays_home_isolation|country),
                  family=Gamma(link=log),
                  glmerControl(optimizer = "bobyqa", optCtrl=list(maxfun=100000)),
                  data=ana_data4) 
summary(control) 

# Gamma mixed-effects accounting for endogeneity of fixed effects
controlendo=glmer(new_outcome~
                    zlog_days+zmdays_country+zmdays_region+
                    zlog_wdays_home_isolation+zmean_hdays_country+zmean_hdays_region+
                    zlog_wdays_business_closure+zmean_bdays_country+zmean_bdays_region+
                    (1+zlog_days+zlog_wdays_home_isolation|region)+
                    (1+zlog_days+zlog_wdays_home_isolation|country),
                  family=Gamma(link=log),
                  glmerControl(optimizer = "bobyqa", optCtrl=list(maxfun=100000)),
                  data=ana_data4) 
summary(controlendo) 

### Warning messages in controlendo due to the cluster means of business closure.
# Without those two, no warnings: see model bla
# However, results are very similar and effect plots do barely differ. Keep controlendo
# for further analyses!
bla=glmer(new_outcome~
                    zlog_days+zmdays_region+zmdays_country+
                    zlog_wdays_home_isolation+zmean_hdays_region+zmean_hdays_country+
                    zlog_wdays_business_closure+
                    (1+zlog_days+zlog_wdays_home_isolation|region)+
                    (1+zlog_days+zlog_wdays_home_isolation|country),
                  family=Gamma(link=log),
                  glmerControl(optimizer = "bobyqa", optCtrl=list(maxfun=100000)),
                  data=ana_data4) 
summary(bla) 

# Check for endogeneity bias
anova(controlendo, control, test="Chisq")


### get significance of random slopes 
keepWarnings <- function(expr) {
  localWarnings <- list()
  value <- withCallingHandlers(expr,
                               warning = function(w) {
                                 localWarnings[[length(localWarnings)+1]] <<- w
                                 invokeRestart("muffleWarning")
                               }
  )
  list(value=value, warnings=localWarnings)
}

# determine RS to remove
vars=c("zlog_days_region", "zlog_wdays_home_isolation_region", 
        "zlog_days_country", "zlog_wdays_home_isolation_country")

to.add=c("(1+zlog_wdays_home_isolation|region)+(1+zlog_days+zlog_wdays_home_isolation|country)", 
         "(1+zlog_days|region)+(1+zlog_days+zlog_wdays_home_isolation|country)", 
         "(1+zlog_days+zlog_wdays_home_isolation|region)+(1+zlog_wdays_home_isolation|country)",
         "(1+zlog_days+zlog_wdays_home_isolation|region)+(1+zlog_days|country)")

all.red.valhf=c()

# loop
for(i in 1:length(to.add)){
  model=paste(c("new_outcome~zlog_days+zmdays_country+zmdays_region+
                zlog_wdays_home_isolation+zmean_hdays_country+zmean_hdays_region+
                zlog_wdays_business_closure+zmean_bdays_country+zmean_bdays_region", 
                  to.add[i]), collapse="+")
  model=as.formula(model)
  red=keepWarnings(glmer(model,
                         family=Gamma(link=log),
                         glmerControl(optimizer = "bobyqa", optCtrl=list(maxfun=100000)),
                         data=ana_data4))
  all.red.valhf=rbind(all.red.valhf, 
                      data.frame(as.data.frame(anova(red$value, controlendo, 
                                  test="Chisq"))[2, c("Chisq", "Df", "Pr(>Chisq)")], 
                                  n.warns=length(unlist(red$warnigs))))
}

all.red.valhf$term=vars
rownames(all.red.valhf)=all.red.valhf$term
pvals_rs=round(all.red.valhf[, 1:3], 4)
pvals_rs[, 3]=pvals_rs[, 3]/2


### get significance of fixed effects
keepWarnings <- function(expr) {
  localWarnings <- list()
  value <- withCallingHandlers(expr,
                               warning = function(w) {
                                 localWarnings[[length(localWarnings)+1]] <<- w
                                 invokeRestart("muffleWarning")
                               }
  )
  list(value=value, warnings=localWarnings)
}

# determine FE to remove
to.rem=c("zlog_days", "zmdays_country", "zmdays_region", "zlog_wdays_home_isolation", 
         "zmean_hdays_country", "zmean_hdays_region", "zlog_wdays_business_closure", 
         "zmean_bdays_country", "zmean_bdays_region")
all.red.valhf=c()

# loop
for(i in 1:length(to.rem)){
  model=paste(c(to.rem[-i]), collapse="+")
  model=paste("new_outcome", model, sep="~")
  model=paste(model,  "(1+zlog_days+zlog_wdays_home_isolation|region)+
              (1+zlog_days+zlog_wdays_home_isolation|country)", sep="+")
  model=as.formula(model)
  red=keepWarnings(glmer(model,
                         family=Gamma(link=log),
                         glmerControl(optimizer = "bobyqa", optCtrl=list(maxfun=100000)),
                         data=ana_data4))
  all.red.valhf=rbind(all.red.valhf, 
                      data.frame(as.data.frame(anova(red$value, controlendo, test="Chisq"))[2, c("Chisq", "Df", "Pr(>Chisq)")], 
                                 n.warns=length(unlist(red$warnigs))))
}

all.red.valhf$term=to.rem
rownames(all.red.valhf)=all.red.valhf$term
pvals_fe=round(all.red.valhf[, 1:3], 4)


# Sensitivity analysis
# Variables
variables=c("(Intercept)", "zlog_days", "zmdays_country", "zmdays_region", 
            "zlog_wdays_home_isolation", "zmean_hdays_country", "zmean_hdays_region",
            "zlog_wdays_business_closure", "zmean_bdays_country", "zmean_bdays_region")

# result matrix 
ori=summary(controlendo)$coefficients[, "Estimate"]
countries=unique(as.character(ana_data4$country))
res_sensanalysis_countries=as.data.frame(matrix(nrow=length(countries), ncol=length(variables)))
colnames(res_sensanalysis_countries)=variables
convergence=rep(NA, times=10)

# loop
for(i in 1:length(countries)){
  test.data=ana_data4[! as.character(ana_data4$country) %in% countries[i], ]
  full=glmer(new_outcome~
               zlog_days+zmdays_country+zmdays_region+
               zlog_wdays_home_isolation+zmean_hdays_country+zmean_hdays_region+
               zlog_wdays_business_closure+zmean_bdays_country+zmean_bdays_region+
               (1+zlog_days+zlog_wdays_home_isolation|region)+
               (1+zlog_days+zlog_wdays_home_isolation|country),
               family=Gamma(link=log),
               glmerControl(optimizer = "bobyqa", optCtrl=list(maxfun=100000)),
               data=test.data)
  coeft=summary(full)$coefficients[, "Estimate"]
  res_sensanalysis_countries[i, ]=coeft
  convergence[i]=any(grepl("failed to converge", full@optinfo$conv$lme4$messages)) 
}

# summary
sens_countries=apply(res_sensanalysis_countries, 2, range) 
sens_range_country=cbind(sens_countries[1, ], ori, sens_countries[2, ]) 
colnames(sens_range_country)=c("min", "original", "max")
sens_range_country 

# Repeat for models that did not converge (2 models)
again=which(convergence==TRUE)

for(i in 1:length(again)){
  sel=again[i]
  test.data=ana_data4[! as.character(ana_data4$country) %in% countries[sel], ]
  full=glmer(new_outcome~
               zlog_days+zmdays_country+zmdays_region+
               zlog_wdays_home_isolation+zmean_hdays_country+zmean_hdays_region+
               zlog_wdays_business_closure+zmean_bdays_country+zmean_bdays_region+
               (1+zlog_days+zlog_wdays_home_isolation|region)+
               (1+zlog_days+zlog_wdays_home_isolation|country),
               family=Gamma(link=log),
               glmerControl(optimizer = "nloptwrap", optCtrl=list(maxfun=100000)),
               data=test.data)
  coeft=summary(full)$coefficients[, "Estimate"]
  res_sensanalysis_countries[sel, ]=coeft
  convergence[sel]=any(grepl("failed to converge", full@optinfo$conv$lme4$messages)) 
}

# summary
sens_countries=apply(res_sensanalysis_countries, 2, range) 
sens_range_country=cbind(sens_countries[1, ], ori, sens_countries[2, ]) 
colnames(sens_range_country)=c("min", "original", "max")
sens_range_country 


### check for multicollinearity
linear=lm(new_outcome~zlog_days+zmdays_country+zmdays_region+
                      zlog_wdays_home_isolation+zmean_hdays_country+zmean_hdays_region+
                      zlog_wdays_business_closure+zmean_bdays_country+zmean_bdays_region,
                      data=ana_data4) 

# car package needed
library(car)
vif(linear)

linear2=lm(new_outcome~zlog_days+zlog_wdays_home_isolation+
          zlog_wdays_business_closure,
          data=ana_data4) 
vif(linear2)


### distribution of random terms
windows()
par(mfrow=c(2, 3))
hist(ranef(controlendo)$region[, "(Intercept)"], main="Region: RI")
hist(ranef(controlendo)$region[, "zlog_wdays_home_isolation"], main="Region: RS home")
hist(ranef(controlendo)$region[, "zlog_days"], main="Region: RS days")
hist(ranef(controlendo)$country[, "(Intercept)"], main="Country: RI")
hist(ranef(controlendo)$country[, "zlog_wdays_home_isolation"], main="Country: RS home")
hist(ranef(controlendo)$country[, "zlog_days"], main="Country: RS days")


### Check for overdispersion
# Function for overdispersion test was provided by Roger Mundry
# load function
source(paste(folder, "/overdisp.R", sep=""))
overdisp=overdisp.test(controlendo)


#### bootstrap f�r confidence intervalls 
# Function for bootstrapping was provided by Roger Mundry
source(paste(folder, "/boot_glmm.R", sep=""))
ci=boot.glmm.pred(control,nboots=1000, use.u=TRUE) # Caution: this might take a while!!!


### Plot effect for entire populations (countries and sub-national units combined)
red=adjustcolor(col="red", alpha.f=0.5)
blue=adjustcolor(col="blue", alpha.f=0.5)
orange=adjustcolor(col="orange", alpha.f=0.5)

windows(6.7, 5)

nf <- layout(matrix(c(1, 1, 1, 1, 2, 2, 2, 2,
                      1, 1, 1, 1, 2, 2, 2, 2,
                      1, 1, 1, 1, 2, 2, 2, 2,
                      1, 1, 1, 1, 2, 2, 2, 2,
                      1, 1, 1, 1, 2, 2, 2, 2,
                      3, 3, 3, 3, 3, 3, 3, 3),
                    6, 8,byrow = TRUE), respect=TRUE)

layout.show(nf)

par(mar=c(2.5,2.5,0.1,0.6), mgp=c(1.2, 0.05, 0), tcl=-0.1, las=1, 
    family="sans")

### First subplot
# Observed data
plot(ana_data4$days, ana_data4$diff_log_cum_confirmed_cases, 
     main= "", xlab="Day", ylab="Growth rate",
     pch=19, col=grey(level=0.5, alpha=0.5))

mtext("All countries", side=3, line=-1.3, font=2)

# Get xvals for predictions 
xvals=seq(min(ana_data4$zlog_days), 
          max(ana_data4$zlog_days), length.out=100)

# Transform xvals back into sequence of days
xvalst=exp(xvals*sd(ana_data4$log_days)+mean(ana_data4$log_days))

# Transform sequence of days into sequence of weighted days with home isolation 
# scale(log(x+1)) 
xvalsh=log(xvalst+1)-mean(log(ana_data4$wdays_home_isolation+1))/
  sd(log(ana_data4$wdays_home_isolation+1))

# 95% CI Fitted vals control
ci.fittedc=lapply(1:nrow(ci$all.boots), function(x){
    ci$all.boots[x, "(Intercept)"]+ 
    ci$all.boots[x, "zlog_days"]*xvals
})
ci.fittedc=matrix(unlist(ci.fittedc), ncol=length(ci.fittedc), byrow=F)
ci.fittedc=t(apply(ci.fittedc, 1, quantile, na.rm=TRUE, probs=c(0.025, 0.975)))
ci.fittedc=exp(ci.fittedc)

# Model prediction
fittedc=exp(xvals*(summary(controlendo)$coefficients["zlog_days", "Estimate"])+
              summary(controlendo)$coefficients["(Intercept)", "Estimate"])

# 95% CI Fitted vals home isolation
ci.fittedh=lapply(1:nrow(ci$all.boots), function(x){
    ci$all.boots[x, "(Intercept)"] + 
    (ci$all.boots[x, "zlog_days"])*xvals +
    (ci$all.boots[x, "zlog_wdays_home_isolation"])*xvalsh
})
ci.fittedh=matrix(unlist(ci.fittedh), ncol=length(ci.fittedh), byrow=F)
ci.fittedh=t(apply(ci.fittedh, 1, quantile, na.rm=TRUE, probs=c(0.025, 0.975)))
ci.fittedh=exp(ci.fittedh)

# model predictions
fittedh=exp(xvals*(summary(controlendo)$coefficients["zlog_days", "Estimate"])+
              xvalsh*(summary(controlendo)$coefficients["zlog_wdays_home_isolation", "Estimate"])+
              summary(controlendo)$coefficients["(Intercept)", "Estimate"])

# Add predictions into plot
polygon(c(xvalst, rev(xvalst)), c(ci.fittedc[, 1], rev(ci.fittedc[, 2])), col=blue, border=NA)
polygon(c(xvalst, rev(xvalst)), c(ci.fittedh[, 1], rev(ci.fittedh[, 2])), col=red, border=NA)
lines(xvalst, fittedc, col="blue", lwd=2, lty=2)
lines(xvalst, fittedh, col="red", lwd=2, lty=3)

### Second subplot
# plot data
plot(ana_data4$days, ana_data4$diff_log_cum_confirmed_cases, 
     main= "", xlab="Day", ylab="Growth rate",
     pch=19, col=grey(level=0.5, alpha=0.5))

mtext("All countries", side=3, line=-1.3, font=2)

# transform sequence of days in sequence of weighted days with business closure
# scale(log(x+1)) 
xvalsb=log(xvalst+1)-mean(log(ana_data4$wdays_business_closure+1))/
  sd(log(ana_data4$wdays_business_closure+1))

# 95% CI Fitted vals home isolation + business closure
ci.fittedb=lapply(1:nrow(ci$all.boots), function(x){
    ci$all.boots[x, "(Intercept)"] + 
    (ci$all.boots[x, "zlog_days"])*xvals +
    (ci$all.boots[x, "zlog_wdays_home_isolation"])*xvalsh+
    (ci$all.boots[x, "zlog_wdays_business_closure"])*xvalsb
})
ci.fittedb=matrix(unlist(ci.fittedb), ncol=length(ci.fittedb), byrow=F)
ci.fittedb=t(apply(ci.fittedb, 1, quantile, na.rm=TRUE, probs=c(0.025, 0.975)))
ci.fittedb=exp(ci.fittedb)

# Model predictions
fittedb=exp(xvals*(summary(controlendo)$coefficients["zlog_days", "Estimate"])+
              xvalsh*(summary(controlendo)$coefficients["zlog_wdays_home_isolation", "Estimate"])+
              xvalsb*(summary(controlendo)$coefficients["zlog_wdays_business_closure", "Estimate"])+
              summary(controlendo)$coefficients["(Intercept)", "Estimate"])

# Add predictions into subplot
polygon(c(xvalst, rev(xvalst)), c(ci.fittedc[, 1], rev(ci.fittedc[, 2])), col=blue, border=NA)
polygon(c(xvalst, rev(xvalst)), c(ci.fittedb[, 1], rev(ci.fittedb[, 2])), col=orange, border=NA)
lines(xvalst, fittedc, col="blue", lwd=2, lty=2)
lines(xvalst, fittedb, col="orange", lwd=2, lty=4)

# Legend
extent<-matrix(ncol=2, nrow=2)
extent[1, 1]<- 1 
extent[1, 2]<- 1
extent[2, 1]<- 5
extent[2, 2]<- 4

par(mar=c(0.3,0.3,0.3,0.3))
plot(extent, type="n", xaxt="n", yaxt="n") 

lines(c(0.9, 1.4), c(3.25, 3.25), lty=2, lwd=2, col=blue)
text(1.5, 3, "Counterfactual: no lockdown (95% CI)", adj=0, font=2, cex=1)
lines(c(0.9, 1.4), c(2.75, 2.75), lty=1, lwd=6, col=blue)

lines(c(0.9, 1.4), c(1.75, 1.75), lty=3, lwd=2, col=red)
text(1.5, 1.5, "Counterfactual: home isolation (95% CI)", adj=0, font=2, cex=1)
lines(c(0.9, 1.4), c(1.25, 1.25), lty=1, lwd=6, col=red)

lines(c(3.2, 3.7), c(3.25, 3.25), lty=4, lwd=2, col=orange)
text(3.8, 3.25, "Counterfactual: home isolation", adj=0, font=2, cex=1)
lines(c(3.2, 3.7), c(2.75, 2.75), lty=1, lwd=6, col=orange)
text(3.8, 2.75, "and business closure (95% CI)", adj=0, font=2, cex=1) 

points(4, 1.5, pch=19, col="grey")
text(4, 1.5, "Observed", pos=4)


### Plot country effect
countries=c("Corea", "Sweden", "UK", "France", "Germany", "Iran", "Italy", 
            "Netherland", "Spain", "USA")
titles=c("South Korea", "Sweden", "England", "France", "Germany", "Iran", "Italy", 
         "Netherlands", "Spain", "USA")

ylabs=rep(c("Growth rate", ""), times=5)
xlabs=c(rep("", times=8), "Day", "Day")

marginsleft=c(1,2.3,0.1,0.2)
marginsright=c(1,1.2,0.1,0.3)
marginsbleft=c(2.3,2.3,0.1,0.3)
marginsbright=c(2.3,1.2,0.1,0.3)
margins=rbind(marginsleft,
              marginsright,
              marginsleft,
              marginsright,
              marginsleft,
              marginsright,
              marginsleft,
              marginsright,
              marginsbleft,
              marginsbright)

blue=adjustcolor(col="blue", alpha.f=0.4)
red=adjustcolor(col="red", alpha.f=0.4)

windows(18, 25)

nf <- layout(matrix(c(1, 1, 1, 1, 1, 1, 1, 1, 2, 2, 2, 2, 2, 2, 2, 2,
                      1, 1, 1, 1, 1, 1, 1, 1, 2, 2, 2, 2, 2, 2, 2, 2, 
                      1, 1, 1, 1, 1, 1, 1, 1, 2, 2, 2, 2, 2, 2, 2, 2, 
                      1, 1, 1, 1, 1, 1, 1, 1, 2, 2, 2, 2, 2, 2, 2, 2, 
                      3, 3, 3, 3, 3, 3, 3, 3, 4, 4, 4, 4, 4, 4, 4, 4,
                      3, 3, 3, 3, 3, 3, 3, 3, 4, 4, 4, 4, 4, 4, 4, 4,
                      3, 3, 3, 3, 3, 3, 3, 3, 4, 4, 4, 4, 4, 4, 4, 4,                      
                      3, 3, 3, 3, 3, 3, 3, 3, 4, 4, 4, 4, 4, 4, 4, 4,
                      5, 5, 5, 5, 5, 5, 5, 5, 6, 6, 6, 6, 6, 6, 6, 6,
                      5, 5, 5, 5, 5, 5, 5, 5, 6, 6, 6, 6, 6, 6, 6, 6,
                      5, 5, 5, 5, 5, 5, 5, 5, 6, 6, 6, 6, 6, 6, 6, 6,
                      5, 5, 5, 5, 5, 5, 5, 5, 6, 6, 6, 6, 6, 6, 6, 6,
                      7, 7, 7, 7, 7, 7, 7, 7, 8, 8, 8, 8, 8, 8, 8, 8, 
                      7, 7, 7, 7, 7, 7, 7, 7, 8, 8, 8, 8, 8, 8, 8, 8, 
                      7, 7, 7, 7, 7, 7, 7, 7, 8, 8, 8, 8, 8, 8, 8, 8, 
                      7, 7, 7, 7, 7, 7, 7, 7, 8, 8, 8, 8, 8, 8, 8, 8, 
                      9, 9, 9, 9, 9, 9, 9, 9, 10, 10, 10, 10, 10, 10, 10, 10,
                      9, 9, 9, 9, 9, 9, 9, 9, 10, 10, 10, 10, 10, 10, 10, 10,
                      9, 9, 9, 9, 9, 9, 9, 9, 10, 10, 10, 10, 10, 10, 10, 10,
                      9, 9, 9, 9, 9, 9, 9, 9, 10, 10, 10, 10, 10, 10, 10, 10,
                      11, 11, 11, 11, 11, 11, 11, 11, 11, 11, 11, 11, 11, 11, 11, 11, 
                      11, 11, 11, 11, 11, 11, 11, 11, 11, 11, 11, 11, 11, 11, 11, 11),
                    22, 16,byrow = TRUE), respect=TRUE)

layout.show(nf)

for(i in 1:length(countries)){
  par(mar=margins[i, ], mgp=c(1.2, 0.05, 0), tcl=-0.1, las=1, 
      family="sans")
  plot(ana_data4$days[ana_data4$country==countries[i]], 
       ana_data4$diff_log_cum_confirmed_cases[ana_data4$country==countries[i]], 
       main= "", xlab=xlabs[i], ylab=ylabs[i],
       pch=19, col=grey(level=0.5, alpha=0.7))
  mtext(titles[i], side=3, line=-1.3, font=2)

  # xvals for predictions
  xvals=seq(min(ana_data4$zlog_days[ana_data4$country==countries[i]]), 
            max(ana_data4$zlog_days[ana_data4$country==countries[i]]), length.out=100)

  # transform xvals into sequence of days
  xvalst=exp(xvals*sd(ana_data4$log_days)+mean(ana_data4$log_days))

  # transform sequence of days into sequence of weighted days with home isolation
  # scale(log(x+1))
  xvalsh=log(xvalst+1)-mean(log(ana_data4$wdays_home_isolation+1))/
  sd(log(ana_data4$wdays_home_isolation+1))

  # 95% CI Fitted vals control
  ci.fittedc=lapply(1:nrow(ci$all.boots), function(x){
      ci$all.boots[x, "(Intercept)"] + 
      ci$all.boots[x, paste("country@(Intercept)@", countries[i], sep="")] +
      (ci$all.boots[x, "zlog_days"]+
      ci$all.boots[x, paste("country@zlog_days@", countries[i], sep="")])*xvals +
      ci$all.boots[x, "zmdays_country"]*
      unique(ana_data4$zmdays_country[ana_data4$country==countries[i]])+
      ci$all.boots[x, "zmean_bdays_country"]*
      unique(ana_data4$zmean_bdays_country[ana_data4$country==countries[i]])
  })
  ci.fittedc=matrix(unlist(ci.fittedc), ncol=length(ci.fittedc), byrow=F)
  ci.fittedc=t(apply(ci.fittedc, 1, quantile, na.rm=TRUE, probs=c(0.025, 0.975)))
  ci.fittedc=exp(ci.fittedc)
  
  # Model predictions control
  fittedc=exp(xvals*(summary(controlendo)$coefficients["zlog_days", "Estimate"]+
                ranef(controlendo)$country[countries[i], "zlog_days"])+
                summary(controlendo)$coefficients["(Intercept)", "Estimate"]+
                ranef(controlendo)$country[countries[i], "(Intercept)"]+
                summary(controlendo)$coefficients["zmdays_country", "Estimate"]*
                unique(ana_data4$zmdays_country[ana_data4$country==countries[i]])+
                summary(controlendo)$coefficients["zmean_bdays_country", "Estimate"]*
                unique(ana_data4$zmean_bdays_country[ana_data4$country==countries[i]]))
  
  ### 95% CI Fitted vals treatment
  ci.fittedt=lapply(1:nrow(ci$all.boots), function(x){
      ci$all.boots[x, "(Intercept)"] + 
      ci$all.boots[x, paste("country@(Intercept)@", countries[i], sep="")] +
      (ci$all.boots[x, "zlog_days"]+
      ci$all.boots[x, paste("country@zlog_days@", countries[i], sep="")])*xvals +
      (ci$all.boots[x, "zlog_wdays_home_isolation"]+
      ci$all.boots[x, paste("country@zlog_wdays_home_isolation@", countries[i], sep="")])*xvalsh+
      ci$all.boots[x, "zmdays_country"]*
      unique(ana_data4$zmdays_country[ana_data4$country==countries[i]])+
      ci$all.boots[x, "zmean_hdays_country"]*
      unique(ana_data4$zmean_hdays_country[ana_data4$country==countries[i]])+
      ci$all.boots[x, "zmean_bdays_country"]*
      unique(ana_data4$zmean_bdays_country[ana_data4$country==countries[i]])
  })
  ci.fittedt=matrix(unlist(ci.fittedt), ncol=length(ci.fittedt), byrow=F)
  ci.fittedt=t(apply(ci.fittedt, 1, quantile, na.rm=TRUE, probs=c(0.025, 0.975)))
  ci.fittedt=exp(ci.fittedt)
  
  # Model predictions
  fittedt=exp(xvals*(summary(controlendo)$coefficients["zlog_days", "Estimate"]+
                ranef(controlendo)$country[countries[i], "zlog_days"])+
                xvalsh*(summary(controlendo)$coefficients["zlog_wdays_home_isolation", "Estimate"]+
                ranef(controlendo)$country[countries[i], "zlog_wdays_home_isolation"])+
                summary(controlendo)$coefficients["(Intercept)", "Estimate"]+
                ranef(controlendo)$country[countries[i], "(Intercept)"]+
                summary(controlendo)$coefficients["zmdays_country", "Estimate"]*
                unique(ana_data4$zmdays_country[ana_data4$country==countries[i]])+
                summary(controlendo)$coefficients["zmean_hdays_country", "Estimate"]*
                unique(ana_data4$zmean_hdays_country[ana_data4$country==countries[i]])+
                summary(controlendo)$coefficients["zmean_bdays_country", "Estimate"]*
                unique(ana_data4$zmean_bdays_country[ana_data4$country==countries[i]]))

  # Add predictions to plot
  polygon(c(xvalst, rev(xvalst)), c(ci.fittedc[, 1], rev(ci.fittedc[, 2])), col=blue, border=NA)
  polygon(c(xvalst, rev(xvalst)), c(ci.fittedt[, 1], rev(ci.fittedt[, 2])), col=red, border=NA)
  lines(xvalst, fittedc, col="blue", lwd=2, lty=2)
  lines(xvalst, fittedt, col="red", lwd=2, lty=3)

  # Get median day of implementation and add to plot
  reg=unique(ana_data4$region)[grepl(countries[i], unique(ana_data4$region))]  
  imp=c()
  for(k in 1:length(reg)){
    check=ana_data4[ana_data4$region==reg[k],]
    cont=min(check$days_home_isolation[check$days_home_isolation>0])
    imp=c(imp, check$days[check$days_home_isolation==cont])
  }
  abline(v=median(imp))
}
  
# Legend
extent<-matrix(ncol=2, nrow=2)
extent[1, 1]<- 1 
extent[1, 2]<- 1
extent[2, 1]<- 5
extent[2, 2]<- 4

par(mar=c(0.1,0.3,0.1,0.3))
plot(extent, type="n", xaxt="n", yaxt="n") 

lines(c(1, 1.5), c(3.25, 3.25), lty=2, lwd=2, col=blue)
text(1.6, 3, "Counterfactual: no home isolation (95% CI)", adj=0, font=2, cex=1)
lines(c(1, 1.5), c(2.75, 2.75), lty=1, lwd=6, col=blue)

lines(c(1, 1.5), c(1.75, 1.75), lty=3, lwd=2, col=red)
text(1.6, 1.5, "Counterfactual: home isolation (95% CI)", adj=0, font=2, cex=1)
lines(c(1, 1.5), c(1.25, 1.25), lty=1, lwd=6, col=red)

points(4, 3, pch=19, col="grey")
text(4, 3, "Observed", pos=4)

lines(c(4, 4), c(1.2, 2.4))
text(4, 2, "Median day of", pos=4)
text(4, 1.4, "implementation", pos=4)


### Plot sub-national unit effect
# Plot 10 strongest deviation from mean for Spain
# get 5 strongest positive and negative effects for Spain
bladata=ranef(controlendo)$region
bladata[order(bladata$zlog_wdays_home_isolation), ]

countries=c("Spain Cataluña", "Spain La Rioja", "Spain Castilla-La Mancha", "Spain Navarra", 
            "Spain Castilla y León",  "Spain Extremadura", "Spain Andalucía", "Spain Murcia",         
            "Spain Cantabria", "Spain Galicia")

titles=c("Catalonia", "La Rioja", "Castilla-La Mancha", "Navarra", 
         "Castilla and Le�n",  "Extremadura", "Andalusia", "Murcia",             
         "Cantabria", "Galicia")

ylabs=rep(c("Growth rate", ""), times=5)
xlabs=c(rep("", times=8), "Day", "Day")

marginsleft=c(1,2.3,0.1,0.2)
marginsright=c(1,1.2,0.1,0.3)
marginsbleft=c(2.3,2.3,0.1,0.3)
marginsbright=c(2.3,1.2,0.1,0.3)
margins=rbind(marginsleft,
              marginsright,
              marginsleft,
              marginsright,
              marginsleft,
              marginsright,
              marginsleft,
              marginsright,
              marginsbleft,
              marginsbright)

blue=adjustcolor(col="blue", alpha.f=0.4)
red=adjustcolor(col="red", alpha.f=0.4)

windows(18, 25)

nf <- layout(matrix(c(1, 1, 1, 1, 1, 1, 1, 1, 2, 2, 2, 2, 2, 2, 2, 2,
                      1, 1, 1, 1, 1, 1, 1, 1, 2, 2, 2, 2, 2, 2, 2, 2, 
                      1, 1, 1, 1, 1, 1, 1, 1, 2, 2, 2, 2, 2, 2, 2, 2, 
                      1, 1, 1, 1, 1, 1, 1, 1, 2, 2, 2, 2, 2, 2, 2, 2, 
                      3, 3, 3, 3, 3, 3, 3, 3, 4, 4, 4, 4, 4, 4, 4, 4,
                      3, 3, 3, 3, 3, 3, 3, 3, 4, 4, 4, 4, 4, 4, 4, 4,
                      3, 3, 3, 3, 3, 3, 3, 3, 4, 4, 4, 4, 4, 4, 4, 4,                      
                      3, 3, 3, 3, 3, 3, 3, 3, 4, 4, 4, 4, 4, 4, 4, 4,
                      5, 5, 5, 5, 5, 5, 5, 5, 6, 6, 6, 6, 6, 6, 6, 6,
                      5, 5, 5, 5, 5, 5, 5, 5, 6, 6, 6, 6, 6, 6, 6, 6,
                      5, 5, 5, 5, 5, 5, 5, 5, 6, 6, 6, 6, 6, 6, 6, 6,
                      5, 5, 5, 5, 5, 5, 5, 5, 6, 6, 6, 6, 6, 6, 6, 6,
                      7, 7, 7, 7, 7, 7, 7, 7, 8, 8, 8, 8, 8, 8, 8, 8, 
                      7, 7, 7, 7, 7, 7, 7, 7, 8, 8, 8, 8, 8, 8, 8, 8, 
                      7, 7, 7, 7, 7, 7, 7, 7, 8, 8, 8, 8, 8, 8, 8, 8, 
                      7, 7, 7, 7, 7, 7, 7, 7, 8, 8, 8, 8, 8, 8, 8, 8, 
                      9, 9, 9, 9, 9, 9, 9, 9, 10, 10, 10, 10, 10, 10, 10, 10,
                      9, 9, 9, 9, 9, 9, 9, 9, 10, 10, 10, 10, 10, 10, 10, 10,
                      9, 9, 9, 9, 9, 9, 9, 9, 10, 10, 10, 10, 10, 10, 10, 10,
                      9, 9, 9, 9, 9, 9, 9, 9, 10, 10, 10, 10, 10, 10, 10, 10,
                      11, 11, 11, 11, 11, 11, 11, 11, 11, 11, 11, 11, 11, 11, 11, 11, 
                      11, 11, 11, 11, 11, 11, 11, 11, 11, 11, 11, 11, 11, 11, 11, 11),
                    22, 16,byrow = TRUE), respect=TRUE)

layout.show(nf)

for(i in 1:length(countries)){
  par(mar=margins[i, ], mgp=c(1.2, 0.05, 0), tcl=-0.1, las=1, 
      family="sans")
  plot(ana_data4$days[ana_data4$region==countries[i]], 
       ana_data4$diff_log_cum_confirmed_cases[ana_data4$region==countries[i]], 
       main= "", xlab=xlabs[i], ylab=ylabs[i],
       pch=19, col=grey(level=0.5, alpha=0.7))
  mtext(titles[i], side=3, line=-1.3, font=2)
  
  # xvals for model predictions
  xvals=seq(min(ana_data4$zlog_days[ana_data4$region==countries[i]]), 
            max(ana_data4$zlog_days[ana_data4$region==countries[i]]), length.out=100)
  
  # transform xvals into sequence of days
  xvalst=exp(xvals*sd(ana_data4$log_days)+mean(ana_data4$log_days))

  # transform sequen of days into sequence of weighted days with home isolation
  # scale(log(x+1))
  xvalsh=log(xvalst+1)-mean(log(ana_data4$wdays_home_isolation+1))/
    sd(log(ana_data4$wdays_home_isolation+1))

  ### 95% CI Fitted vals control
  ci.fittedc=lapply(1:nrow(ci$all.boots), function(x){
      ci$all.boots[x, "(Intercept)"] + 
      ci$all.boots[x, "country@(Intercept)@Spain"] +
      ci$all.boots[x, paste("region@(Intercept)@", countries[i], sep="")] +
      (ci$all.boots[x, "zlog_days"]+
      ci$all.boots[x, "country@zlog_days@Spain"]+
      ci$all.boots[x, paste("region@zlog_days@", countries[i], sep="")])*xvals+
      ci$all.boots[x, "zmdays_country"]*
      unique(ana_data4$zmdays_country[ana_data4$country=="Spain"])+
      ci$all.boots[x, "zmdays_region"]*
      unique(ana_data4$zmdays_region[ana_data4$region==countries[i]])+
      ci$all.boots[x, "zmean_bdays_country"]*
      unique(ana_data4$zmean_bdays_country[ana_data4$country=="Spain"])+
      ci$all.boots[x, "zmean_bdays_region"]*
      unique(ana_data4$zmean_bdays_region[ana_data4$region==countries[i]])
  })
  ci.fittedc=matrix(unlist(ci.fittedc), ncol=length(ci.fittedc), byrow=F)
  ci.fittedc=t(apply(ci.fittedc, 1, quantile, na.rm=TRUE, probs=c(0.025, 0.975)))
  ci.fittedc=exp(ci.fittedc)
  
  # Model predictions
  fittedc=exp(xvals*(summary(controlendo)$coefficients["zlog_days", "Estimate"]+
                       ranef(controlendo)$country["Spain", "zlog_days"]+
                       ranef(controlendo)$region[countries[i], "zlog_days"])+
                       summary(controlendo)$coefficients["(Intercept)", "Estimate"]+
                       ranef(controlendo)$country["Spain", "(Intercept)"]+
                       ranef(controlendo)$region[countries[i], "(Intercept)"]+
                       summary(controlendo)$coefficients["zmdays_country", "Estimate"]*
                       unique(ana_data4$zmdays_country[ana_data4$country=="Spain"])+
                       summary(controlendo)$coefficients["zmdays_region", "Estimate"]*
                       unique(ana_data4$zmdays_region[ana_data4$region==countries[i]])+
                       summary(controlendo)$coefficients["zmean_bdays_country", "Estimate"]*
                       unique(ana_data4$zmean_bdays_country[ana_data4$country=="Spain"])+
                       summary(controlendo)$coefficients["zmean_bdays_region", "Estimate"]*
                       unique(ana_data4$zmean_bdays_region[ana_data4$region==countries[i]]))
  
  ### 95% CI Fitted vals treatment
  ci.fittedt=lapply(1:nrow(ci$all.boots), function(x){
      ci$all.boots[x, "(Intercept)"] + 
      ci$all.boots[x, "country@(Intercept)@Spain"] + 
      ci$all.boots[x, paste("region@(Intercept)@", countries[i], sep="")] +
      (ci$all.boots[x, "zlog_days"]+ 
      ci$all.boots[x, "country@zlog_days@Spain"] +
      ci$all.boots[x, paste("region@zlog_days@", countries[i], sep="")])*xvals +
      (ci$all.boots[x, "zlog_wdays_home_isolation"]+ 
      ci$all.boots[x, "country@zlog_wdays_home_isolation@Spain"] +
      ci$all.boots[x, paste("region@zlog_wdays_home_isolation@", countries[i], sep="")])*xvalsh+
      ci$all.boots[x, "zmdays_country"]*
      unique(ana_data4$zmdays_country[ana_data4$country=="Spain"])+
      ci$all.boots[x, "zmdays_region"]*
      unique(ana_data4$zmdays_region[ana_data4$region==countries[i]])+
      ci$all.boots[x, "zmean_bdays_country"]*
      unique(ana_data4$zmean_bdays_country[ana_data4$country=="Spain"])+
      ci$all.boots[x, "zmean_bdays_region"]*
      unique(ana_data4$zmean_bdays_region[ana_data4$region==countries[i]])+
      ci$all.boots[x, "zmean_hdays_country"]*
      unique(ana_data4$zmean_hdays_country[ana_data4$country=="Spain"])+
      ci$all.boots[x, "zmean_hdays_region"]*
      unique(ana_data4$zmean_hdays_region[ana_data4$region==countries[i]])
  })
  ci.fittedt=matrix(unlist(ci.fittedt), ncol=length(ci.fittedt), byrow=F)
  ci.fittedt=t(apply(ci.fittedt, 1, quantile, na.rm=TRUE, probs=c(0.025, 0.975)))
  ci.fittedt=exp(ci.fittedt)

  # Model predictions  
  fittedt=exp(xvals*(summary(controlendo)$coefficients["zlog_days", "Estimate"]+
                ranef(controlendo)$country["Spain", "zlog_days"]+
                ranef(controlendo)$region[countries[i], "zlog_days"])+
                xvalsh*(summary(controlendo)$coefficients["zlog_wdays_home_isolation", "Estimate"]+
                ranef(controlendo)$country["Spain", "zlog_wdays_home_isolation"]+
                ranef(controlendo)$region[countries[i], "zlog_wdays_home_isolation"])+
                summary(controlendo)$coefficients["(Intercept)", "Estimate"]+
                ranef(controlendo)$country["Spain", "(Intercept)"]+
                ranef(controlendo)$region[countries[i], "(Intercept)"]+
                summary(controlendo)$coefficients["zmdays_country", "Estimate"]*
                unique(ana_data4$zmdays_country[ana_data4$country=="Spain"])+
                summary(controlendo)$coefficients["zmdays_region", "Estimate"]*
                unique(ana_data4$zmdays_region[ana_data4$region==countries[i]])+
                summary(controlendo)$coefficients["zmean_bdays_country", "Estimate"]*
                unique(ana_data4$zmean_bdays_country[ana_data4$country=="Spain"])+
                summary(controlendo)$coefficients["zmean_bdays_region", "Estimate"]*
                unique(ana_data4$zmean_bdays_region[ana_data4$region==countries[i]])+
                summary(controlendo)$coefficients["zmean_hdays_country", "Estimate"]*
                unique(ana_data4$zmean_hdays_country[ana_data4$country=="Spain"])+
                summary(controlendo)$coefficients["zmean_hdays_region", "Estimate"]*
                unique(ana_data4$zmean_hdays_region[ana_data4$region==countries[i]]))
  
  # Add predictions to plot
  polygon(c(xvalst, rev(xvalst)), c(ci.fittedc[, 1], rev(ci.fittedc[, 2])), col=blue, border=NA)
  polygon(c(xvalst, rev(xvalst)), c(ci.fittedt[, 1], rev(ci.fittedt[, 2])), col=red, border=NA)
  lines(xvalst, fittedc, col="blue", lwd=2, lty=2)
  lines(xvalst, fittedt, col="red", lwd=2, lty=3)
  
  # Get day of implementation and add to plot
  reg=unique(ana_data4$region)[grepl(countries[i], unique(ana_data4$region))]  
  check=ana_data4[ana_data4$region==reg,]
  cont=min(check$days_home_isolation[check$days_home_isolation>0])
  imp=check$days[check$days_home_isolation==cont]
  abline(v=median(imp))

}

# Legend
extent<-matrix(ncol=2, nrow=2)
extent[1, 1]<- 1 
extent[1, 2]<- 1
extent[2, 1]<- 5
extent[2, 2]<- 4

par(mar=c(0.1,0.3,0.1,0.3))
plot(extent, type="n", xaxt="n", yaxt="n") 

lines(c(1, 1.5), c(3.25, 3.25), lty=2, lwd=2, col=blue)
text(1.6, 3, "Counterfactual: no home isolation (95% CI)", adj=0, font=2, cex=1)
lines(c(1, 1.5), c(2.75, 2.75), lty=1, lwd=6, col=blue)

lines(c(1, 1.5), c(1.75, 1.75), lty=3, lwd=2, col=red)
text(1.6, 1.5, "Counterfactual: home isolation (95% CI)", adj=0, font=2, cex=1)
lines(c(1, 1.5), c(1.25, 1.25), lty=1, lwd=6, col=red)

points(4, 3, pch=19, col="grey")
text(4, 3, "Observed", pos=4)

lines(c(4, 4), c(1.2, 2.4))
text(4, 2, "Day of", pos=4)
text(4, 1.4, "implementation", pos=4)
```
